# Systematic Review and Meta-analysis of Mental Health Impact on BAME Populations with Preterm Birth

**DOI:** 10.1101/2022.03.22.22272780

**Authors:** G Delanerolle, Y Zeng, P Phiri, T Phan, N Tempest, P Busuulwa, A Shetty, V Raymont, S Rathod, JQ Shi, DK Hapangama

## Abstract

**Background:** Preterm birth (PTB) is one of the main causes of neonatal deaths globally, with approximately 15 million infants are born preterm. Therefore, the mental health (MH) impact experienced by mothers experiencing a PTB is important, especially within the Black, Asian and Minority Ethnic (BAME) populations.

**Aim:** The aims of the study were to determine the prevalence of MH conditions among BAME women with PTB as well as the MH assessments used to characterise the MH outcomes.

**Methods:** A systematic methodology was developed and published as a protocol in PROSPERO (CRD42020210863). Multiple databases were used to extract relevant data. I^2^ and Egger’s tests were used to detect the heterogeneity and publication bias. A *Trim and fill* method was used to demonstrate the influence of publication bias and the credibility of conclusions.

**Results:** Thirty-nine studies met the eligibility criteria from a possible 3526. The prevalence rates of depression among PTB-BAME mothers were significantly higher than full-term mothers with a standard median deviation (SMD) of 1.5 and a 95% confidence interval (CI) 29-74%. The subgroup analysis indicated, depressive symptoms to be time sensitive. Women within the *very PTB* category demonstrated a significantly higher prevalence of depression than those categorised as *non-very* PTB. The prevalence rates of anxiety and stress among PTB-BAME mothers were significantly higher than full-term mothers (OR of 88% and 60% with a CI of 42%-149% and 24-106%, respectively).

**Conclusion:** BAME women with PTB suffers with MH conditions. Many studies did not report on BAME population specific MH outcomes. Therefore, the impact of PTB is not accurately represented in this population, and thus could negatively influence the quality of maternity services.

**Core Tip:** This study demonstrates the mental health impact due to preterm birth among the Black, Asian and Ethnic minority women. There is minimal research available at present around this subject matter, and potential disease sequalae.

## 1. Introduction

Preterm birth (PTB) is a multi-etiological condition and a leading cause of perinatal mortality and morbidity [1]. PTB can be categorized as per the World Health Organisation (WHO) classification methods as extreme preterm (gestational age <28 weeks), very preterm (gestational age of 28-32 weeks) and moderately preterm (32-37 weeks). Most preterm infants are at risk of developing respiratory and gastrointestinal complications [2]. The PTB rates are higher in most developed regions of the world despite advances in medicine. PTB are at the highest level in the US for between 12-15% and 5-9% in Europe. In comparison, PTB rates in China range between 4.7-18.9% (1987-2006) and Taiwan 8.2-9.1% [3]. The prevalence of PTB increased from 9.8% in 2000 to 10.6% by 2014 and has become a global public health issue [1]. However, the mental health (MH) impact associated with PTB is not extensively examined, despite it potentially may exacerbate the patient’s experience of a distressing birth. Furthermore, clearly pronounced risk of PTB among Black women have been reported in studies from US or UK [4,5], with limited data on the risk among other ethnic groups. While health disparities, social deprivation are recognised risk factors for PTB that are also frequently associated with Black Asian Minority Ethnic (BAME) populations, the available data on ethnic disparities associated with PTB remains limited.

In the UK alone, health disparities within Caribbean and West African populations demonstrate a significant risk of very PTB in comparison to Caucasians. Similar risks within the South Asian community appear to be less consistent in comparison to Caucasian PTB women [11]. In the UK, National Health Service (NHS) England reports improvements to maternity services are a priority as part of the NHS 10-year plan [12]. As per the 2018 Public Health England report on maternity services, 1 in 4 of all births within Wales and England were to mothers born outside the UK [12]. Additionally, 13% of all infants born between 2013-2017 are from the BAME population [12]. Importantly, Black women were 5 times more at risk of death during parturition and Asian babies are 73% more likely to result in neonatal death compared to Caucasian women [12], therefore, the MH impact experienced by PTB mothers is vital to evaluate particularly in the BAME population. A number of socio-economic, genetic and obstetric causes have been proposed to explain MH disorders among PTB women, but these theories do not fully explain the aetiology. Furthermore, they also exclude the bidirectional relationship between PTB, and MH conditions demonstrated by some studies [13] [18][19].

This available evidence demonstrates a need to explore the MH impact on BAME women with PTB. We believe that gathering this evidence would inform the forthcoming evidence-based women’s health strategy in the UK to explore both the physical and MH components, and to be inclusive using cultural adaptations where appropriate.

## 2. Materials & Methods

An evidence synthesis methodology was developed using a systematic protocol that was developed and published on PROSPERO (CRD42020210863). The aims of the study were to determine the prevalence of MH conditions among BAME women with PTB as well as the MH assessments used to characterise the MH outcomes.

### 2.1 Data searches

Multiple databases were used, including PubMed, EMBASE, Science direct, and The Cochrane Central Register of Controlled trials for the data extraction process. Searches were carried out using multiple keywords and MeSH terms such as *Depression, Anxiety, Mood disorders, PTSD, Psychological distress, Psychological stress, Psychosis, Bipolar, Mental Health, Unipolar, self-harm, BAME, Preterm birth, Maternal wellbeing and Psychiatry disorders*. These terms were then expanded using the ‘snow-ball’ method and the fully developed methods are in the supplementary section (supp. methods 1).

### 2.2 Eligibility criteria and study selection

All eligible randomised controlled trials (RCTs) and non-RCTs published in English were included. The final dataset was reviewed independently. Multiple MH variables were used alongside of the 2 primary variables of PTB and BAME.

### 2.3 Data extraction and Analysis

The extraction and eligibility has been demonstrated using a PRISMA diagram [Supplementary PRISMA Diagram].

The data was collected using Endnote and Microsoft excel. Stata 16.1 was used as a way to complete the final statistical analysis. Standardized Mean Difference (SMD) and 95% Confidence Interval (CI) were extracted for analysis. Heterogeneity was assessed by way of funnel plots, *χ*^2^ -test (P-value) and 1^2^. A sub-group analysis was conducted to determine the MH symptomatologies identified and the geographical location.

Due to the unified use of MH assessments, in order to standardize the mean differences reported within each study, the following mathematical method was used [25-27]:

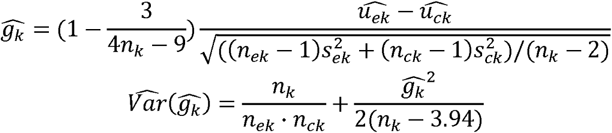

where 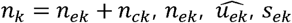 are the number, mean and standard variation of exposed group and 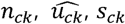 are the number, mean and standard variation of control group. Then we can obtain the 95% confidence interval by

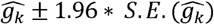

where 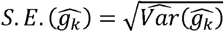.

### 2.4 Meta-regression and sub-group analysis

To eliminate heterogeneity, a meta-regression and sub-group analyses were conducted by MH assessment timepoints and country.

### 2.5 Sensitivity analysis

To further analyse the heterogeneity of studies reporting depression and anxiety, a sensitivity analysis was conducted.

### 2.6 Risk of bias quality assessment

Studies included within this study were critically appraised individually using MH variables. All studies appraised for methodological quality and risk of bias based on the Newcastle-Ottawa-Scale (NOS), which is commonly used for cross-sectional and/or cohort studies as demonstrated by Wells et al. [76]. These could be further modified using the adapted NOS version as reported by Modesti et al. [77] and colleagues. The NOS scale includes 8 items within 3 specific quality parameters of selection, outcome and comparability. The quality of these studies was reported as good, fair or poor based on the details below;

- Good quality score of 3 or 4 stars were awarded in selection, 1 or 2 in comparability and 2 or 3 stars in outcomes
- Fair quality score of 2 stars were awarded in selection, 1 or 2 stars in comparability and 2 or 3 stars in outcomes
- Poor quality score was allocated 0 or 1 star in selection, 0 stars in comparability and 0 or 1 star in outcomes

### 2.7 Outcomes

The following outcomes were included within the meta-analysis;

- Prevalence of anxiety, depression, PTSD and stress
- Clinical significance of the data identified
- Critical interpretive synthesis of common MH reported outcomes

Outcomes such as PPD could not be synthesised for the meta-analysis. Therefore, these aspects have been included in the narrative analysis only.

### 2.8 Publication bias

Publication bias is a concern to the validity of conclusion of a meta-analysis. As a result, several methods could be used to assess this aspect. An egger’s test was used to report on publication bias. Additionally, a *trim and fill* (TAF) method was used to analyse the influence of publication bias. TAF estimates any missing studies due to publication bias within the funnel plot to adjust the overall effect estimate.

### 2.9 Patient and public involvement

A representative from a patient-public focus group associated with a multi-morbid project investigating women’s physical and mental health sequalae was invited to review the protocol and the resulting paper. This is a vital facet of developing and delivering an authentic evidence synthesis to reduce the gap between evidence production, development of solutions to address the identified gaps and the implementation of the solutions into practice as well as their acceptability by patients.

## 3. Results

### 3.1 Meta-analysis

Of the 3526 studies, 39 met the eligibility criteria. All 39 studies reported the MH status of BAME women with PTB although it remained unclear if they reported MH symptoms or clinical diagnoses. Figure 1 shows the PRISMA diagram. The MH assessments and frequency of the data gathering varied across studies. The 39 studies primarily reported stress, anxiety and depression and the key characteristics (Table 1) and the keys features of the MH assessments (Table 2 and Supplementary information) used within these studies are demonstrated in Tables 1 and 2.

**Figure 1:**
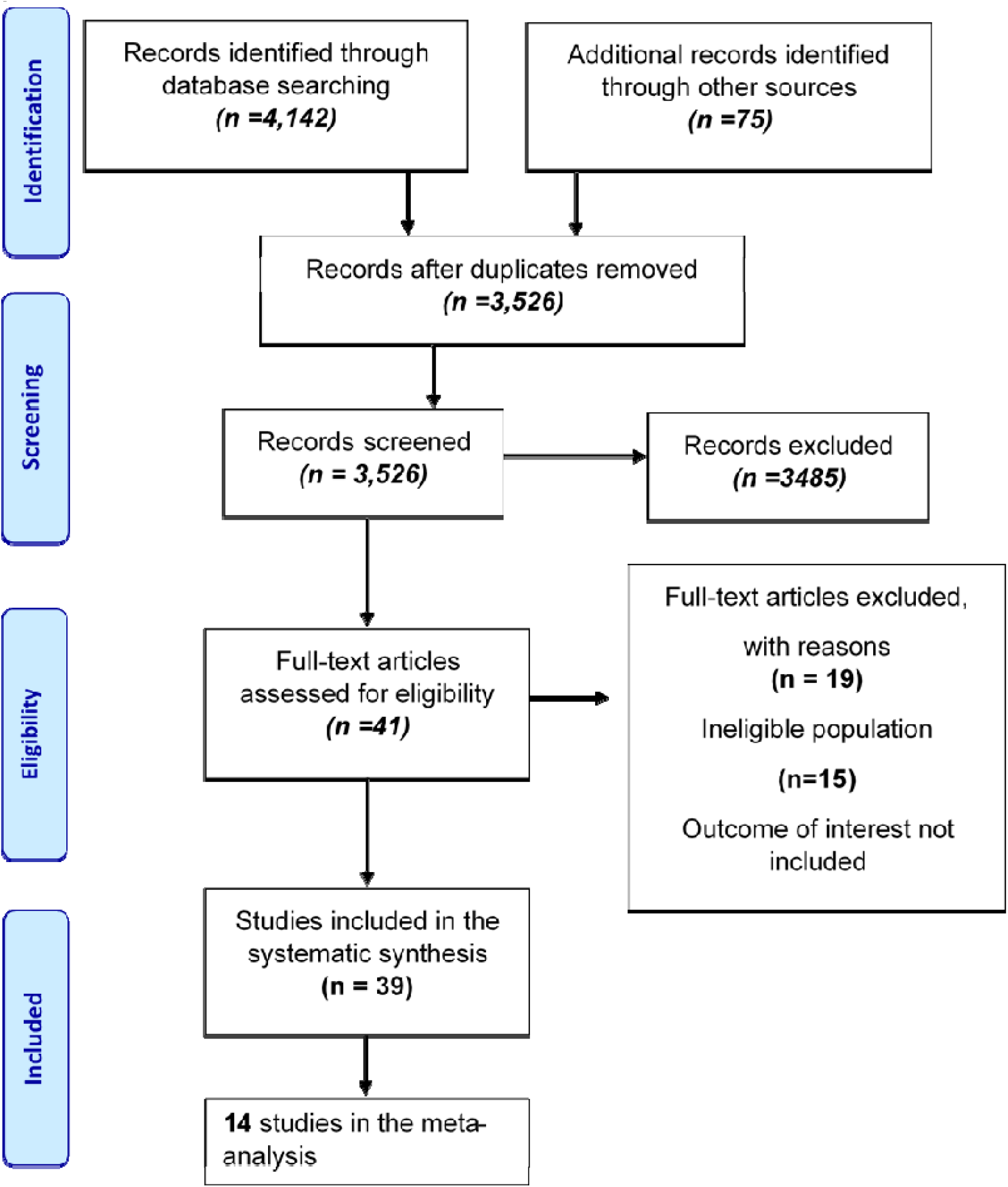
PRISMA Diagram.

**Table 1.**
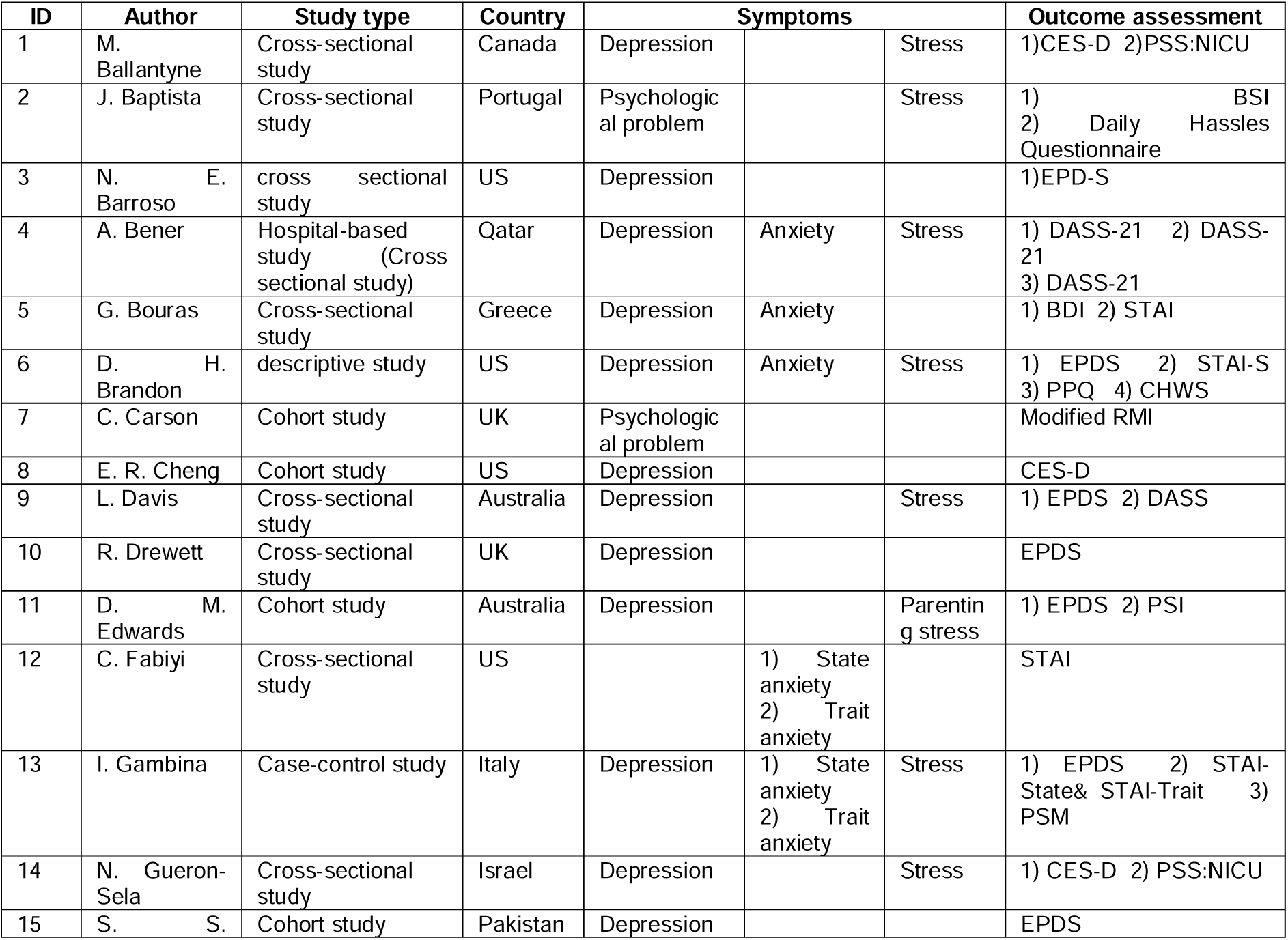
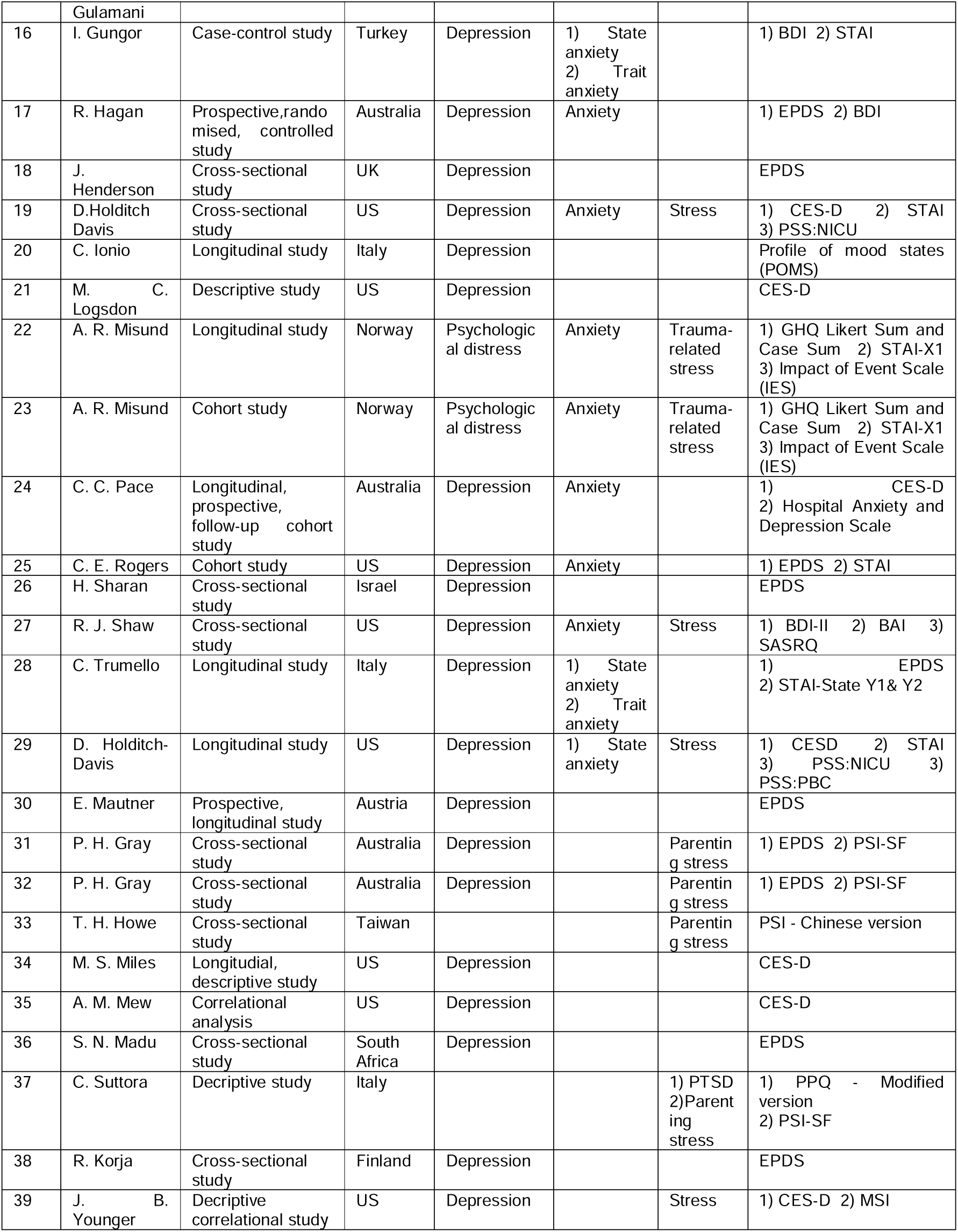
indicate key features of the studies included in the systematic review.

**Table 2.**
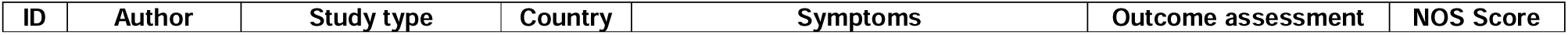
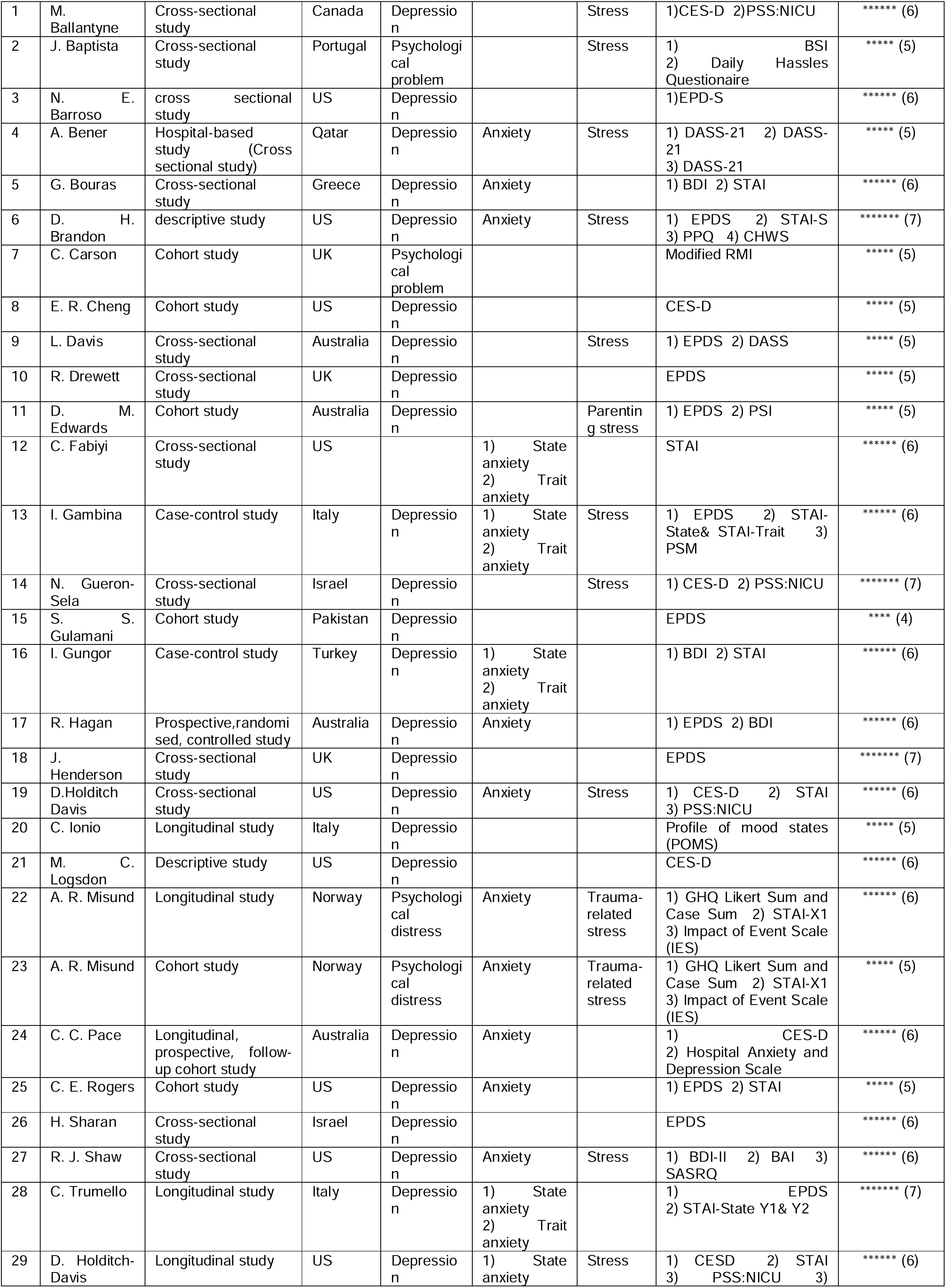
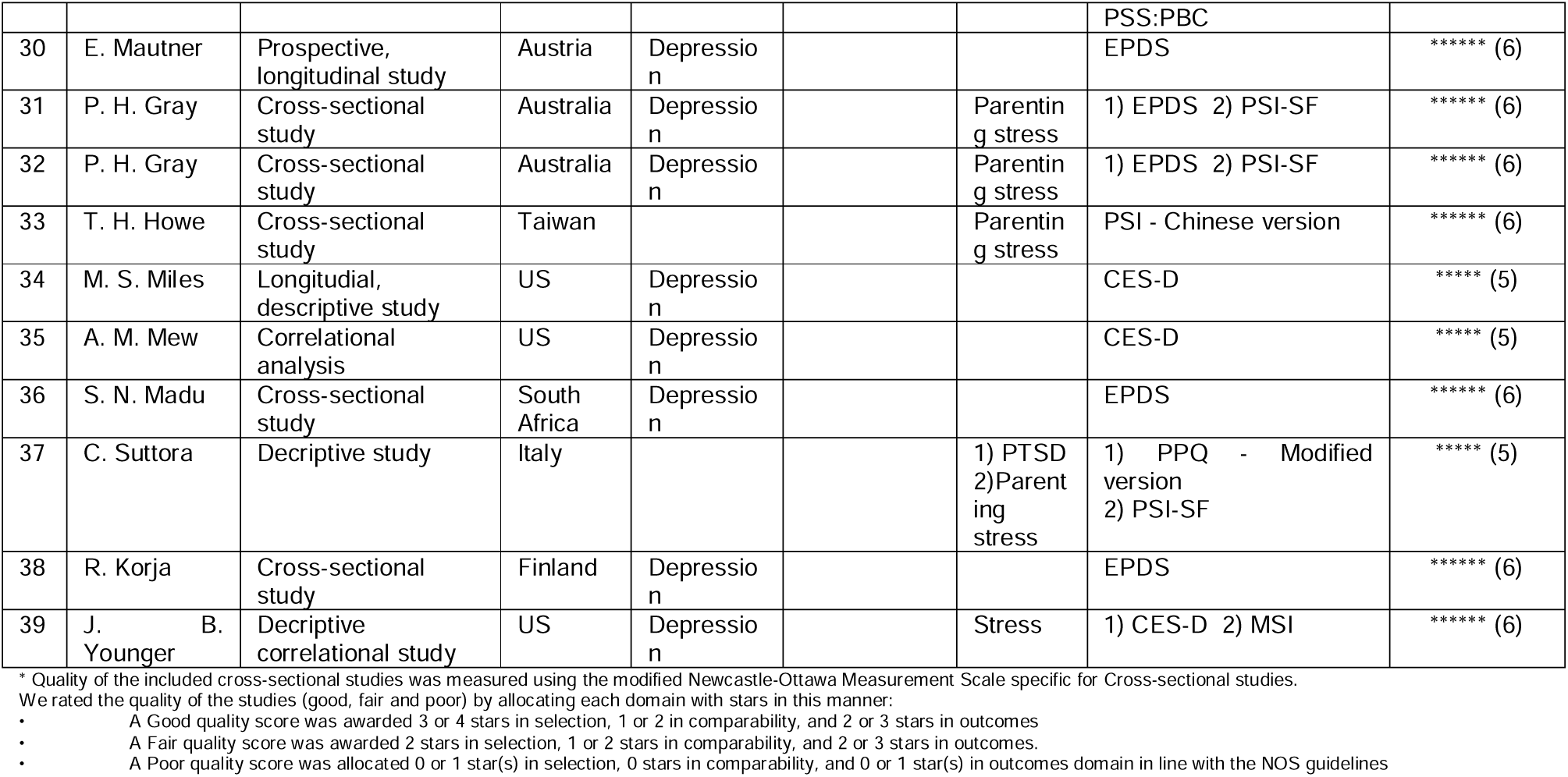
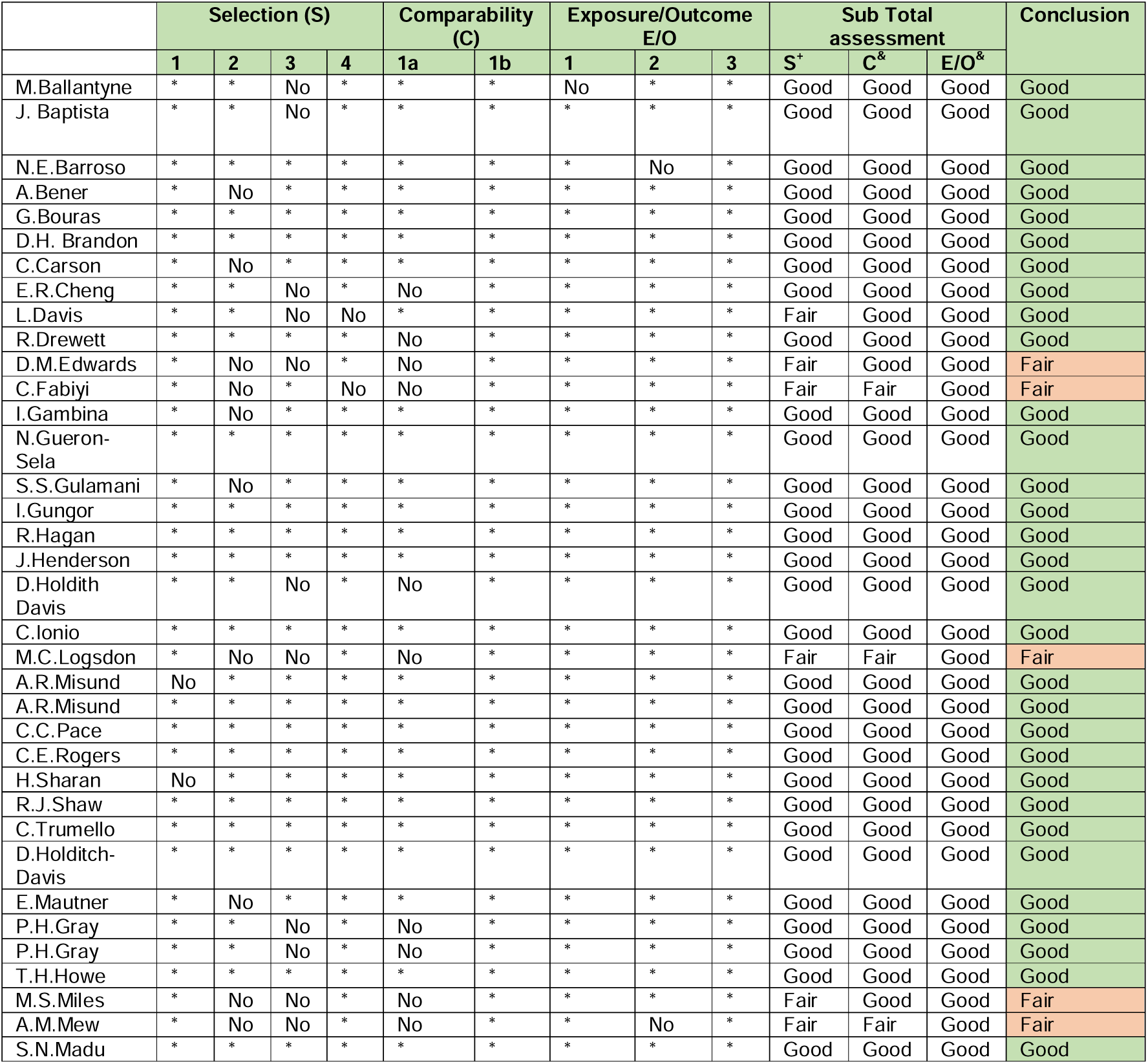
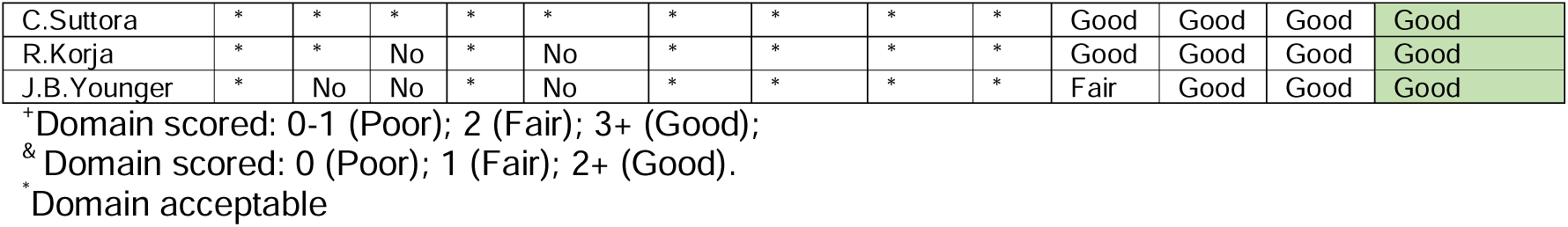
indicate the quality assessment outcomes using the Newcastle-Ottawa Scale.

#### Depression

Of the 39 studies, 36 primarily reported an association between the prevalence of depression and PTB. Fifteen studies only examined the differences of non-depressive symptoms as well other factors such as race, ethnicity, plurality across multiple assessment timepoints although, they were not compared to full-term birth mothers of BAME decent. The overall SMD was 0.4 and 95%CI of a range of 0.25-0.56, indicating the prevalence of depression in PTB mothers to be significantly higher than mothers who delivered at term. *I*^2^=82.69% indicated high heterogeneity among the depression group.

Shaw and colleagues [44] focused on the association between depression symptoms and the efficiency of Edinburgh Postnatal Depression Scale (EPDS), although the specificity of EPDS to the BAME population was not demonstrated. Since most of the studies reported mean and SD, we pooled mean differences and its 95% CI. Seven of the studies lacked information about mean score and SD, thus, were excluded from the meta-analysis. Gray and colleagues [28,29] used the same dataset in two papers, therefore one of these was included into the meta-analysis. Therefore, a total of 12 studies were included in the meta-analysis as indicated by Table 3. Additionally, Gueron-Sela [30] studied two ethnicities, therefore it was used twice as reported in Table 3. Therefore, 13 items were reported in the meta-analysis for depression. The meta-analyses for anxiety and stress had 5 studies each, as demonstrated in Table 4 and 5.

**Table 3.** indicates characteristics of the 12 studies included within the meta-analysis for depression.

| ID | Author | Study type | Country | Sample size | Outcome assessment |
| --- | --- | --- | --- | --- | --- |
| 1 | D. H. Brandon [39] | descriptive study | US | 60 | EPDS |
| 2 | G. Bouras [34] | Cross-sectional study | Greece | 200 | BDI |
| 3 | E. R. Cheng [13] | Cohort study | US | 5350 | CES-D |
| 4 | R. Drewett [51] | Cross-sectional study | UK | 10838 | EPDS |
| 5 | I. Gambina [33] | Case-control study | Italy | 84 | EPDS |
| 6 | P. H. Gray [28] [29] | Cross-sectional study | Australia | 217 | EPDS |
| 7 | N.Gueron-Sela [30] | Cross-sectional study | Israel | 103(Bedouin)<br>230(Jewish) | CES-D |
| 8 | I. Gungor [35] | Case-control study | Turkey | 299 | BDI |
| 9 | C. Ionio [53] | Longitudinal study | Italy | 50 | Profile of mood states (POMS) |
| 10 | S. N. Madu [31] | Cross-sectional study | South Africa | 100 | EPDS |
| 11 | E. Mautner [56] | Prospective, longitudinal study | Australia | 61 | EPDS |
| 12 | C. C. Pace [32] | Longitudinal, prospective cohort study | Australia | 230 | CES-D |

**Table 4.** indicates characteristics of the 5 studies included within the meta-analysis for anxiety.

| ID | Author | Study type | Country | Sample size | Outcome assessment |
| --- | --- | --- | --- | --- | --- |
| 1 | D. H. Brandon [39] | descriptive study | US | 60 | STAI-S |
| 2 | G. Bouras [34] | Cross-sectional study | Greece | 200 | (a) STAI-T<br>(b) STAI-S |
| 3 | I. Gambina [33] | Case-control study | Italy | 84 | (a) STAI-T<br>(b) STAI-S |
| 4 | I. Gungor [35] | Case-control study | Turkey | 299 | (a) STAI-T<br>(b) STAI-S |
| 5 | C. C. Pace [32] | Longitudinal, prospective cohort study | Australia | 230 | HADS-A |

**Table 5.**
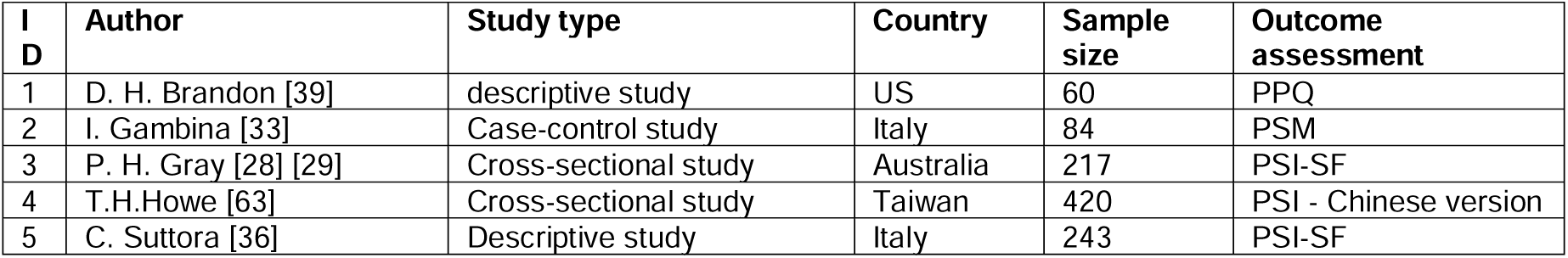
indicates characteristics of the 5 studies included within the meta-analysis for Stress.

#### Anxiety

The 12 studies reporting anxiety utilised EDPS, the State-Trait Anxiety Inventory (STAI), Hospital Anxiety and Depression Scale (HADS-A), Centre for Epidemiological Studies Depression (CES-D), Beck’s Depression Inventory (BDI) and Profile of Mood States (POMS) as their assessment tool. The total scores of these scales are different, and the mean difference of the studies are not compatible. Four studies reported on anxiety using STAI and HADS-A as their MH assessment of choice. The overall SMD of Anxiety was 0.63 with 95%CI of 0.35-0.91. *I*^2^ =86.83% also indicated high heterogeneity among anxiety group.

#### Stress and Parent stress index

Studies reporting stress used the Parent Stress Index (PSI) assessment on three separate timepoints along with the Professional Personality Questionnaire (PPQ) and the Perceived Stress Measure (PSM). The total scores of these scales in each meta-analysis are different, and the mean difference of the studies are not compatible. The overall SMD of Stress was 0.47 with 95%CI 0.22-0.72. *I*^2^=77.55% indicated high heterogeneity among stress group.

#### PTSD

Suttora et al [36] was the only study reporting on PTSD. The reported mean and SD of the symptoms of PTSD were transformed to SMD. The SMD was 1.12 with a 95%CI of 0.84-1.40 indicated significantly high PTSD symptoms among BAME PTB women than the term mothers.

Assessment of MH at differing timepoints for depression, anxiety and stress were evaluated between full term and PTB mothers. Different mean scores and SD values were reported across the included studies. The dataset was unified with converting the mean difference to the standardised mean difference (SMD) and demonstrated in the forest plots (Figures 2, 3 and 4).

**Figure 2:**
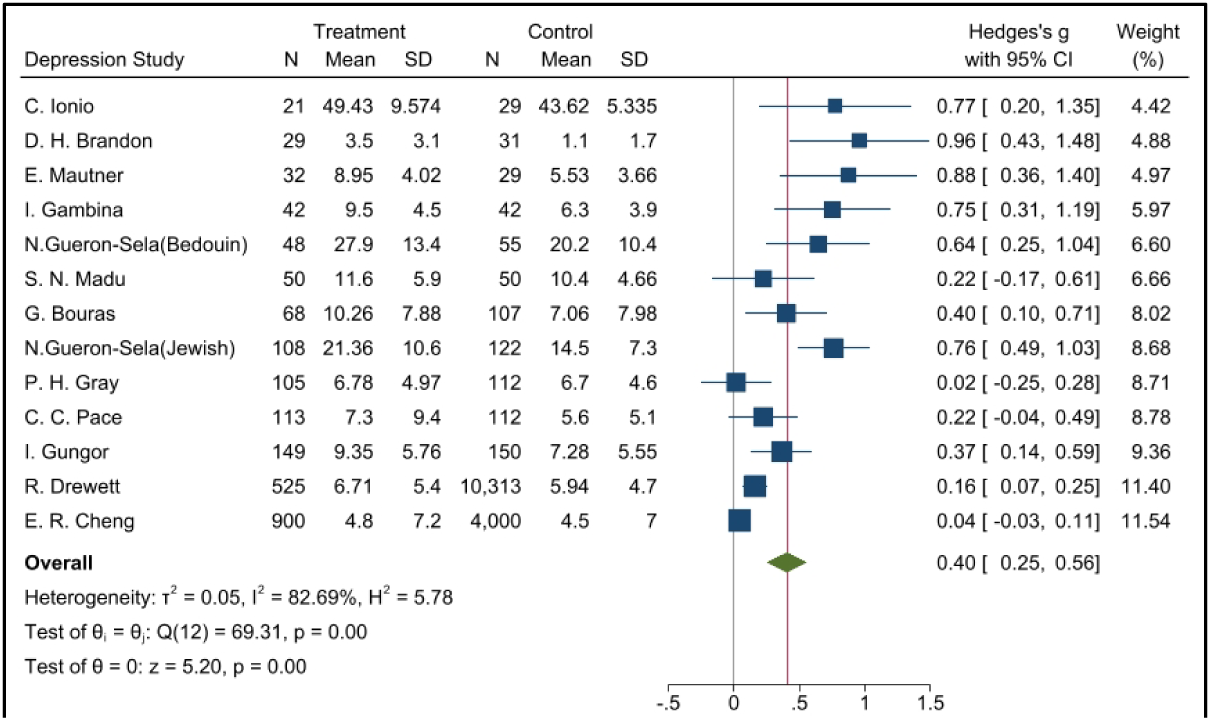
Forest plot for depression (Full term vs. Preterm birth)

**Figure 3:**
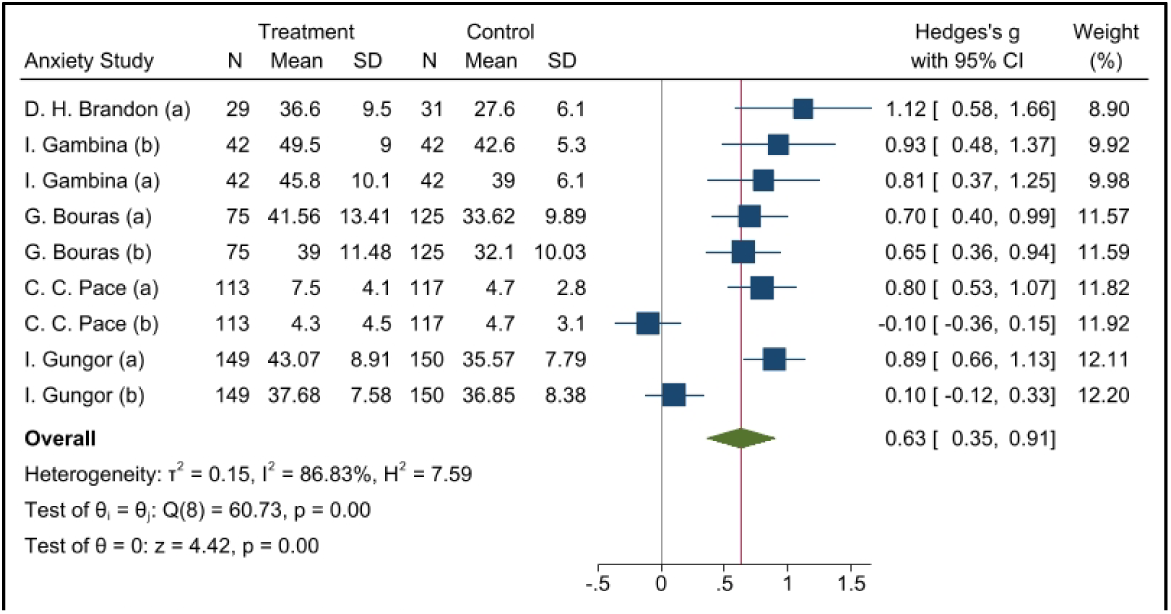
Forest plot for anxiety (Full term vs. Preterm birth)

**Figure 4:**
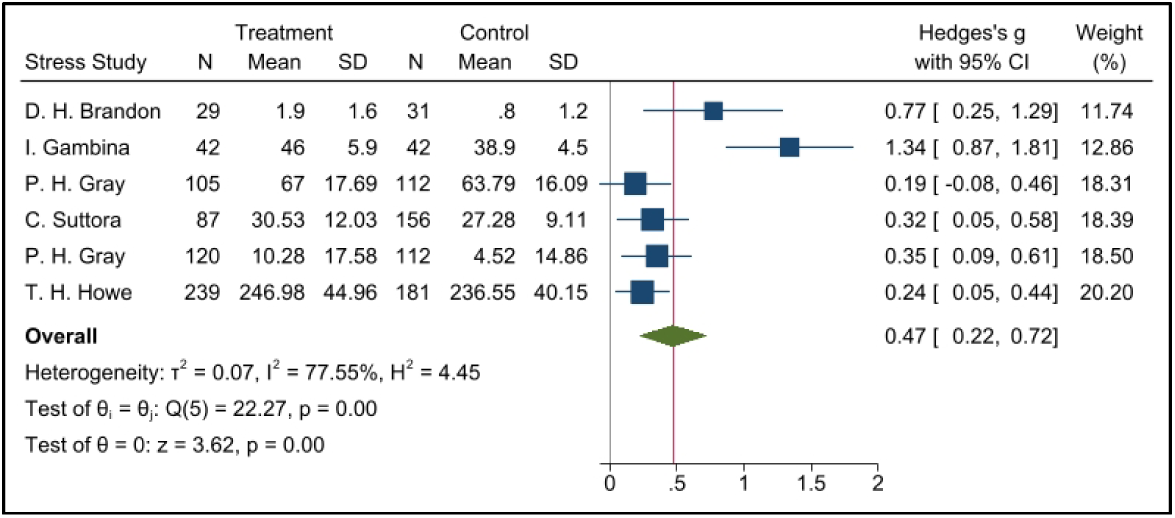
Forest plot for stress (Full term vs. Preterm birth)

This meta-analysis identified depression to be a primary MH outcome among PTB mothers and significantly higher prevalence rates of depression was reported in PTB mothers compared with full-term mothers.

### 3.2 Meta-regression analysis

Of the 16 studies included within the meta-regression analysis for depression, 5 reported mean scores and SD of the MH questionnaires used at parturition. Four studies recorded the mean and SD at 1-month post-delivery, while another four studies reported the same at 1 to 8 months post-delivery. To eliminate the heterogeneity, these studies were adjusted by timepoints (Figure 5).

**Figure 5:**
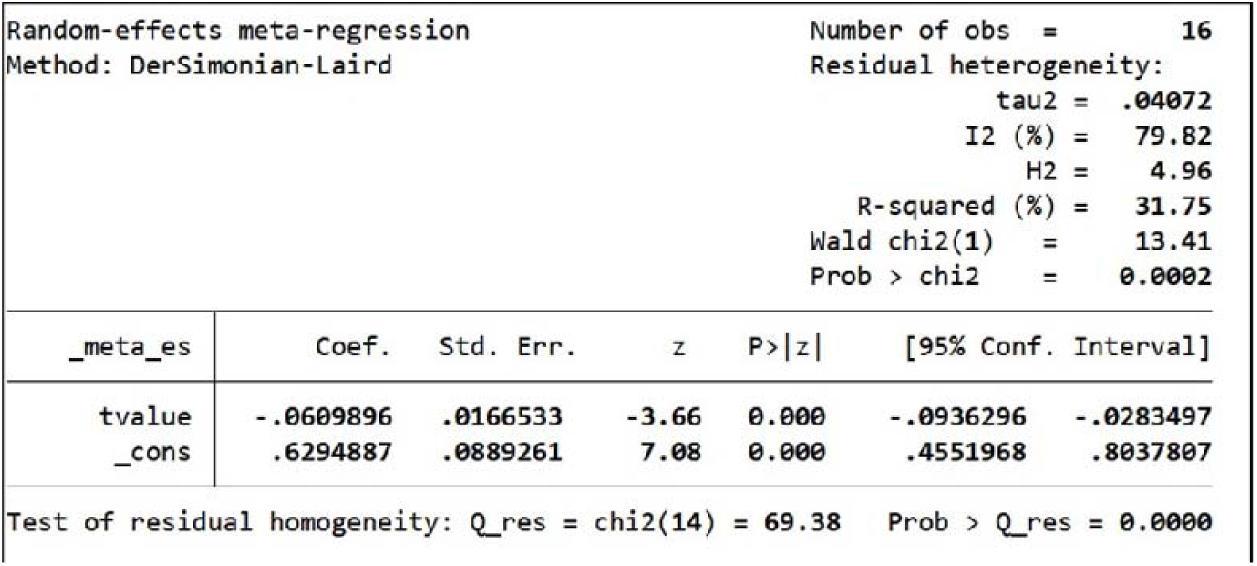
Meta regression conducted by time for depression.

The estimated intercept for depression is 0.629 with a 95% CI of 0.455-0.804. This indicates the MH assessment scores within the PTB group were significantly higher than full-term group at the birth. The coefficient of the covariate time was -0.061 with a 95%CI of -0.094, -0.028 indicating that the coefficients of time were significantly lower than 0. This is indicative of a reduction depression symptoms post-delivery. Heterogeneity decreased from 82.69% to 79.82%, and the differences of assessment timepoints could explain the 31.75% of the heterogeneity identified.

Nine studies that reported anxiety were included in the meta-regression (Figure 6). The estimated intercept was 0.772 with a 95%CI of 0.500-1.045 which indicates the MH assessment scores of the PTB group are significantly higher than the scores of full-term group. The coefficient of the covariate time is -0.136 with 95%CI of -0.262, -0.010 indicating that the symptoms of anxiety gradually disappeared among PTB group following birth. Heterogeneity reduced from 86.83% to 80.29%. The differing timepoints in administering the MH assessment could explain 34.91% of the heterogeneity (Figure 6).

**Figure 6:**
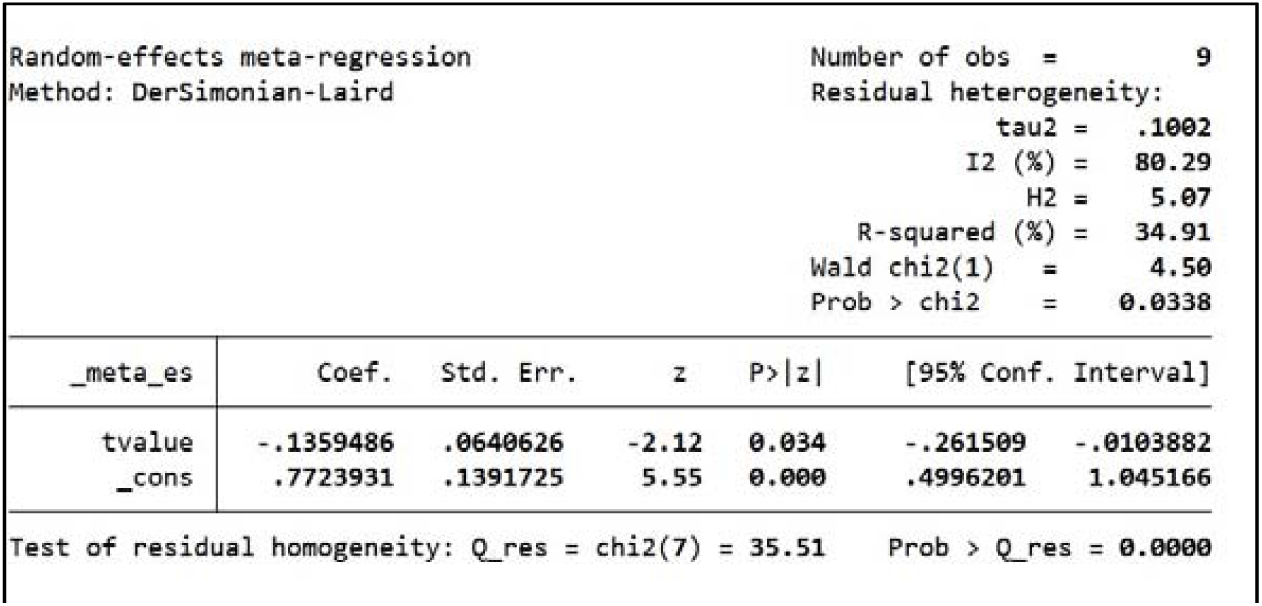
Meta regression conducted by time for anxiety.

Following the reduction of heterogeneity by way of the meta-regression method, the statistical conclusions demonstrate a statistical significance where the prevalence of depression among BAME women with PTB was higher in comparison to BAME women who delivered at full-term. The *I*^2^ was almost 80% which indicates a high heterogeneity.

The pooled SMD within the studies using PTB mothers from USA was 0.46 with a 95%CI of -0.43 – 1.35. The pooled SMD within Australia was 0.44 with a 95% CI of 0.07-0.81. *I*^2^ of these two subgroups indicated a high heterogeneity: 0.38 and 0.18 respectively. The assessment timepoints of these two groups have a significant difference which could be the source of the high heterogeneity. As there were only 2 studies, a meta-regression of the timepoints could not be completed.

### 3.3 Subgroup analysis

A subgroup analysis of depression and anxiety was completed using geographical location as demonstrated in Supplementary Figures 1-3. For depression, the pooled SMD within Greece, Italy, Israel and Turkey was 0.57 with a 95%CI of 0.4-0.74. The pooled SMD within UK was 0.12 with a 95% CI of 0.03-0.21. *I*^2^ was denoted to be indicating a lack of heterogeneity as demonstrated in Supplementary Figure 1.

Although a meta-regression was not conducted for the pooled SMD within the studies with PTB mothers from USA, a subgroup analysis demonstrated that the high heterogeneity could be attributed to the differences of timepoints of the MH assessments.

Of the 39 studies included in the systematic review, thirteen studies were from North America [1,3,6,8,12,19,21,25,27,29,34,35,39], thirteen from Europe [2,5,7,10,13,18,20,22,23,28,30,37,38], six studies from Australia [9,11,17,24,31,32], three from Asia [15,16,33], three from the Middle East [4,14.26] and one from South Africa [36]. These have been demonstrated in Table 1. Depression was the most frequently reported theme across all the studies, followed by anxiety and stress (Table 6). A variety of diagnostic tools were used across the studies, which reflects the diverse clinical practices across different countries.

**Table 6.**
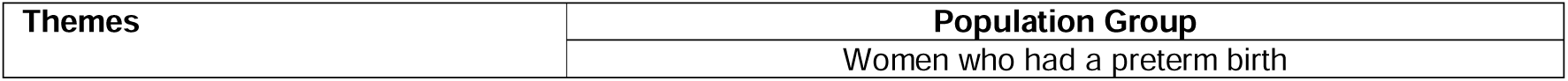
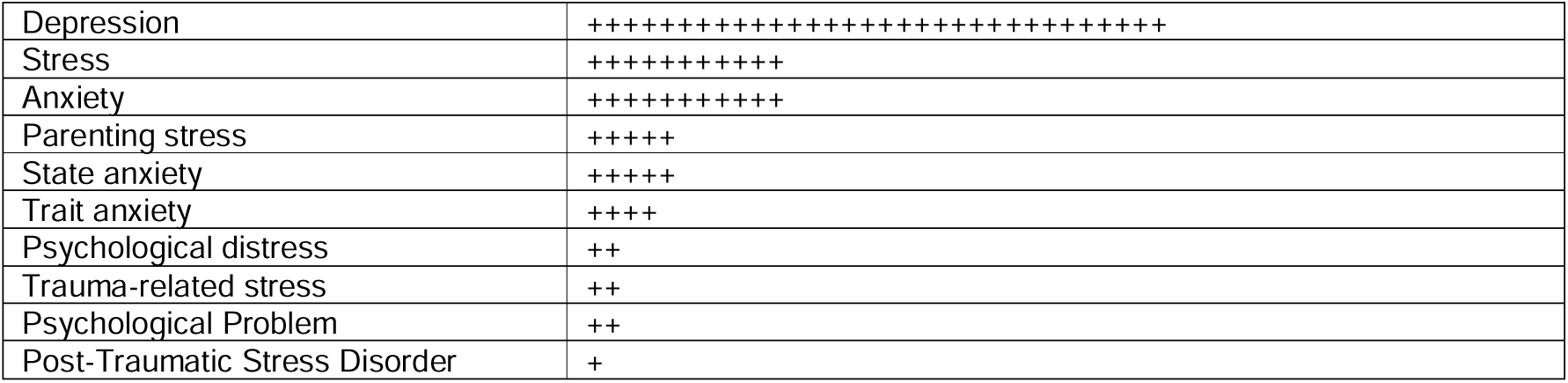
indicates the thematic synthesis.

Based on the identified data, PTB women from the Mediterranean region (Greece, Italy, Turkey and Israel) may be more prone to depressive symptoms in comparison to BAME women with PTB in Australia and the US. The pooled OR and its respective 95% CIs appear credible for PTB BAME women experiencing a significantly higher prevalence of depression post-parturition, although the MH symptoms appear to reduce over time.

The pooled SMD of anxiety within USA was 1.12 with 95%CI of 0.58-1.66 whilst the pooled SMD of the Mediterranean region (Greece, Italy, Turkey) was 0.66 with a 95%CI of 0.37-0.95. The pooled SMD of Australia was 0.35 with a 95%CI of -0.54 -1.23 (Supplementary Figure 2). BAME women with PTB from Australia appear to have less symptoms of anxiety and the main source of the high heterogeneity in subgroup was still from the timepoints.

In relation to assessing stress, Gray and colleagues [28,29] conducted MH assessments at months 4 and 12, post-parturition. Whilst this appears to be useful follow-up data to evaluate, the outcome measures were analysing 2 different MH variables of parenting stress and general stress. As shown in Figure 12, four studies reported on parenting stress and 2 of them reported on the overall state of stress. The subgroup analysis conducted indicated a lack of heterogeneity between these studies. Mild heterogeneity (*I*^2^ = 60.69%) was identified within the studies included in the stress group alone. The pooled SMD within the parenting group was 0.27 with a 95%CI of 0.15-0.39. In the stress group, the pooled SMD was 1.07 with a 95%CI of 0.51-1.62. Additionally, the symptoms of parenting stress were less severe within the PTB group (supplementary figures 4 & 5; Table 6).

### 3.3 Sensitivity analysis

Studies reporting depression [32] demonstrated women with severe PTB indicated a high SMD at parturition indicating elevated levels of depressive symptoms (supplementary Figure 6). A combination of worries about very premature babies and the trauma following parturition may further attribute to elevated depressive symptoms. Only Pace and colleagues’ study [32] conducted the assessment of questionnaires among the very PTB women group at the birth. Women with a more severe PTB may indicate higher scores of depression, therefore this study was excluded from the sensitivity analysis. After removing Pace and colleagues’ study [32], the heterogeneity in Australia reduced from an *I*^2^ of 87.79% to 52.99 %. Therefore, conclusions were adjusted from a pooled SMD of 0.42 (with 95%CI 0.28-0.56) to 0.34 (with 95%CI 0.22-0.46). Despite this numerical change, an elevated level of depression among BAME PTB women were visible in comparison to those with a full-term pregnancy (supplementary Figure 6).

Based on the anxiety studies, Gungor et al [35] in particular, reported extremely small OR and a sensitivity analysis was conducted excluding one possible outlier study, as indicated by supplementary Figure 7. The heterogeneity identified without Gungor et al [35] was 0%. Therefore, this study in particular appears to have design and methodological issues limiting generalisability of the findings. As a result, conclusions were amended from an SMD of 0.63 (with 95%CI 0.35-0.91) to 0.7 (with 95%CI 0.42-0.98). Therefore, despite the amendment [35], a significantly high prevalence among BAME PTB women is observed (supplementary Figure 7).

### 3.4 Publication bias

For studies with a small sample size, the pooled OR is significantly higher based on the funnel plots, therefore, these would be prone to publication bias. To assess this further, Egger’s tests were conducted for all studies included within the meta-analysis.

Funnel plots developed within this sample intuitively revealed publication bias (Supplementary Figures 8-10). Egger’s test of meta-analysis studies for depression (p-value =0.001), indicated the small sample sizes are a source of publication bias (supplementary Figure 11). The pooled SMD 0.4 and associated CI (0.25-0.56) may have been overrated. Therefore, the *trim and fill* method was used to further improve the statistical conclusions (as indicated in supplementary Figures 11 and 12). The asymmetry of the funnel plot demonstrates the studies could minimally impact publication bias.

Based on the findings demonstrated in supplementary Figures 11, 3 further studies were imputed to correct the effect size of small studies. The small study effect was eliminated with using the imputation method, and publication bias was corrected (demonstrated in supplementary Figure 12). The Hedge’s g (supplementary Figure 12) was significantly higher than 0 among the meta-analysis based and imputed studies. After imputing the 3 new studies and removing the publication bias, the statistical conclusion was adjusted from a SMD of 0.4 with 95%CI of 0.25-0.56 to 0.32 (95%CI of 0.18-0.47. Despite the adjustments of publication bias, there was significant evidence that the prevalence of depression among BAME PTB women were higher than those who gave birth at full-term (supplementary Figures 12 and 13).

Egger’s test p-value for anxiety was 0.198, indicating no publication bias exists (demonstrated in supplementary Figure 14).

Egger’s test p-value for stress was 0.036, indicating a slight publication bias among the studies (demonstrated in supplementary Figure 15).

Ascertainment bias was considered within the context of the meta-analysis. Due to the lack of required details such as the proportion of different ethnic groups and MH assessments, it was not possible to assess this numerically. However, within the context of all the studies included in the systematic review portion of the study, it is evident, there could be ascertainment bias as the sampling methods used in the studies comprise of patients who may or may not have a higher or lower probability of reporting MH symptomatologies. These studies may be subjected to selection bias due to the lack of consistency around frequency of administering the relevant MH instruments. In essence, studies should have had samples with all ethnicities and races (including Caucasians) to better evaluate the true MH impact due to PTB. Furthermore, the sample population should have received a standardised set of MH assessments to determine anxiety, depression, PTSD and other mental illnesses at specific time points during the pre and post-natal period since it is common to have undiagnosed MH conditions. In addition to this, some studies have had attempted to evaluate the MH impact after birth at 8 months although this lacks scientific justification and thereby, epidemiologically insignificant. Furthermore, due to the lack of consistency in assessing and reporting MH outcomes postnatally, attrition bias may be present. However, a definitive conclusion could not be attained numerically due to limitations in the sample sizes reported.

## 4 Discussion

In this meta-analysis, the prevalence rate of depression among PTB BAME mothers was identified to be significantly higher than in full-term mothers with an OR of 1.50 and 95%CI of 29%-74%. Depressive symptoms were frequently reported postnatally in mothers and fathers of premature infants [13]. The causes of this are likely multifactorial. Social support may influence this relationship. Cheng et al [13] reported that mothers with non-resident fathers experienced higher rates of depressive symptoms, as did the non-resident fathers included in this study. Lack of social support is likely to be further exacerbated by prolonged hospitalisation of preterm infants and the unique challenges faced by infants following hospital discharge. Additionally, mothers may be admitted to hospital prior to delivery, in some cases for weeks, due to conditions like severe preeclampsia or PROM associated with PTB and hence they may be more isolated than mothers of term infants.

This study defined three sub-groups; assessment timepoint <1 month, 1-8 months and >8 months, and indicated that shorter the time after giving birth, the more significant was the depression. Therefore, the provision of MH support following the immediate post-partum period would benefit patients. Within the first month after delivery, depressive symptoms were significant among PTB mothers; however, by 8 months and after 8 months, the prevalence of depression was only slightly significant among PTB mothers (OR of 1.17 with 95% CI of 8%-27%; OR of 1.06 with a CI of 1%-12%).

Separation of the infant and the mother is an important and frequent occurrence in PTB, which may explain why mothers of preterm infants are at increased risk of depression. Furthermore, maternal co-morbidities including preeclampsia or recovery from an obstetrics intervention such as a caesarean section may also impact on a mother’s ability to bond with her new-born who maybe in a neonatal intensive care (NICU) or special care unit. One study from South Africa [31] demonstrated a high prevalence of depression in mothers of both full term and preterm infants from lower socioeconomic groups, which may have contributed to a greater incidence of depression. Women from lower socioeconomic groups are likely exposed to greater stressors and less resources [31], affecting their MH.

Adjusting to parenthood is important for all parents, in the case of PTB mothers may not have sufficient time to prepare, which may lead to maternal stress [48]. Familiarity with the situation, possibly by having had a previous preterm infant, and predictability have been found to reduce stress and anxiety [48]. Medically indicated preterm delivery is maybe planned, for example, with multiple pregnancies and maternal diabetes and thus, predictable. Therefore, it is possible that those mothers experience less stress than those who give birth following acute spontaneous onset preterm labor. In addition to mental preparation, the former group of parents of preterm infants may have had time to visit the NICU and speak with neonatologists to gain further information and this may reduce anxiety following birth.

Parenting stress is found to be higher in mothers of preterm infants at one year [29]. This relationship may by predicted by maternal depression as well as impaired parent and infant interactions [29]. Interestingly, parenting stress is not significantly different in mothers of preterm or full-term infants in early infancy [28], suggesting all mothers require support in the immediate post-partum period to reduce parental MH but prolonged provision of such support is important in managing PTB mothers.

Increased and unexpected medical interventions associated with PTB, including painful corticosteroid injections or the use of magnesium sulphate, as well as intimate examinations and the need for emergency procedures such as caesarean sections may further negatively impact a mother’s physical and mental health. These may exacerbate the underlying stress faced by a preterm mother and her partner, the feelings of anxiety, stress are compounded in some circumstances by the lack of preparedness and loss of control. Together, these experiences may explain why mothers and fathers of preterm infants have greater levels of stress [29] and depression [13].

Cheng et al [13] conducted the comparison between fathers and mothers suffering as a result of PTB among Hispanic, Non-Hispanic White, Non-Hispanic Black and Non-Hispanic as well as other races. Gueron et al [30] on the other hand focused on depression and stress symptomatologies among Bedouin and Jewish women. Based on Gueron and colleagues’ findings, Bedouin women experienced the highest level of depression. In comparison to these, Rogers et al [43] compared the Caucasian and African American PTB patients that indicated a lack of significant differences between the two groups. Ballantyne et al [37] conducted their study on Canadian PTB women which included immigrant women. However, immigrant’s status had no contribution to the differences in MH disorders or symptomatologies.

The MH impact on those with PTB could be exacerbated due to understandable feelings of helplessness and hopelessness, and low mood is commonly reported by these women. On the contrary, Jotzo and colleagues [14] demonstrated PTB could lead to traumatising effects on parents with 49% of mothers reporting traumatic reactions even after a year. Muller-Nix and colleagues [15] demonstrated this correlation of traumatic stress and psychological distress between mother and child. Pierrehumbert and colleagues [16] indicated post-traumatic stress symptoms after PTB was a predictor of a child’s eating and sleeping problems. Similarly, Solhaugh and colleagues [17] found that parents who had hospital stays following a PTB requiring neonatal intensive care, demonstrated high levels of psychological reactions that required treatment.

Perinatal MH around suicidality or suicidal ideation, should be considered as a priority to be addressed among BAME women, which is vital in particular within the UK. BAME women are at a higher risk of suffering from MH disorder in comparison to Caucasian women in the UK and they are less likely to access healthcare support. This is particularly true for women of Pakistani and Indian background. Additionally, Anderson and colleagues [64] reported prevalence and risk of MH disorders among migrant women. These factors should be considered by those treating clinical groups. In addition to the timepoint, we also consider the impact of population. It remains unclear whether the prevalence rate of depression varies after PTB in different ethnic groups. Gulamani et al [67] have found the depressive symptoms of women with PTB may be associated with race and culture, but further evidence is lacking. Due to the higher risk of MH symptoms around the time of PTB, this data may help the health service providers to focus on delivering timely support to the BAME mothers with PTB.

Interestingly, alcohol consumption and substance abuse that are linked to worsening of MH and poor pregnancy outcomes were not identified within the BAME population literature within the scope of this study, although, it would influence PTB and associated MH illness [68-73].

Similarly, substance abuse among pregnant women increases the risk of PTB and the association of mental illness among the BAME population [74,75]. Holden et al [75] demonstrated self-reported depressive symptomatologies associated with 602 BAME and Caucasian pregnant women that had substance abuse and were subjected to intimate partner violence. This study used the EPDMS which demonstrated elevated levels of depressive illness that required clinical diagnoses and treatments at a MH care facility. Additionally, women abuse screening tool (WAST) was used to evaluate relationship issues and those needing appropriate support was referred to social services [74, 75]. There is limited information available around substance abuse and partner violence associated with MH among BAME women. Research conducted within this area appears to lack consistency and systematic evaluation of cultural paradigms relevant to BAME women and the direct association with PTB and MH given the complexity of these issues.

## 5 Limitations

Heterogeneity of studies gathered within this review challenged the evidence synthesis. Studies identified reported on MH outcomes without a clear distinction mostly between MH symptomatologies and psychiatric comorbidities. Timelines for administering MH instruments and other tools such as talking therapies were not unified across all studies. Collectively, these are design and methodological flaws influencing heterogeneity. Studies were excluded if they discussed QoL as this does not demonstrate the identification or reporting of MH outcomes such as pre or postnatal depression, anxiety, psychosis and other mood disorders.

## 6 Comments & Conclusion

PTB has a significant association with depression, anxiety and stress symptoms in new mothers during the immediate postpartum period. The MH symptoms are more significant in very preterm mothers than non-very preterm mothers. However, the effect of PTB on the incidence of depression and other MH outcomes is unclear among different ethnic groups and therefore more studies are needed to explore this.

This study identified a methodological gap to evaluate disease *sequalae* between PTB and MH among BAME populations. This important facet should be considered in future research studies that requires the involvement of multidisciplinary teams. Most included studies did not indicate a publicly available protocol, and this would have assisted in reducing potential biases during selection to improve sampling techniques and the subsequent data analysis. PICO based reporting may benefit future researchers when conducting research within PTB to address true MH impact within BAME populations. Addressing the multi-stakeholder evidence gap is crucial to improving patient care, therefore, the development of a classification framework for healthcare systems to better assess BAME women at risk with PTB and MH outcomes would be beneficial. Including cultural adaptation methods as well as training of healthcare professionals will help to manage patients’ expectations with the required sensitivities. Similarly, cost-effectiveness and long-term sustainability should be considered when developing a suitable framework.

It is also vital to acknowledge health inequalities and avoidable disparities should be addressed as a matter of urgency. Maternal care should have integrated methods of working with MH care professionals and possibly setup a specialist service for BAME women with a focus on culturally adapted and sensitive ways to support women after a PTB. It is important to improve quality of care received by BAME women with vulnerabilities such as those who are refugees or migrants and do not speak English. Equally, MH services should work more cohesively within the women’s health in the community setting and training should be offered to all healthcare professionals to provide a personalised care.

## Data Availability

All data produced in the present study are available upon reasonable request to the authors

## Acknowledgements

The authors acknowledge support from Southern Health NHS Foundation Trust, University College London and Liverpool Women’s hospital. We would like to acknowledge Mrs Nyla Haque who inspired the discussion of BAME groups within the context of this study.

This paper is part of the multifaceted ELEMI project that is sponsored by Southern Health NHS Foundation Trust and in collaboration with the University of Liverpool, Liverpool Women’s Hospital, University College London, University College London NHS Foundation Trust, University of Southampton, Robinson Institute-University of Adelaide, Ramaiah Memorial Hospital (India), University of Geneva and Manchester University NHS Foundation Trust.

## Supplementary Figures

**Supplementary Figure 1.**
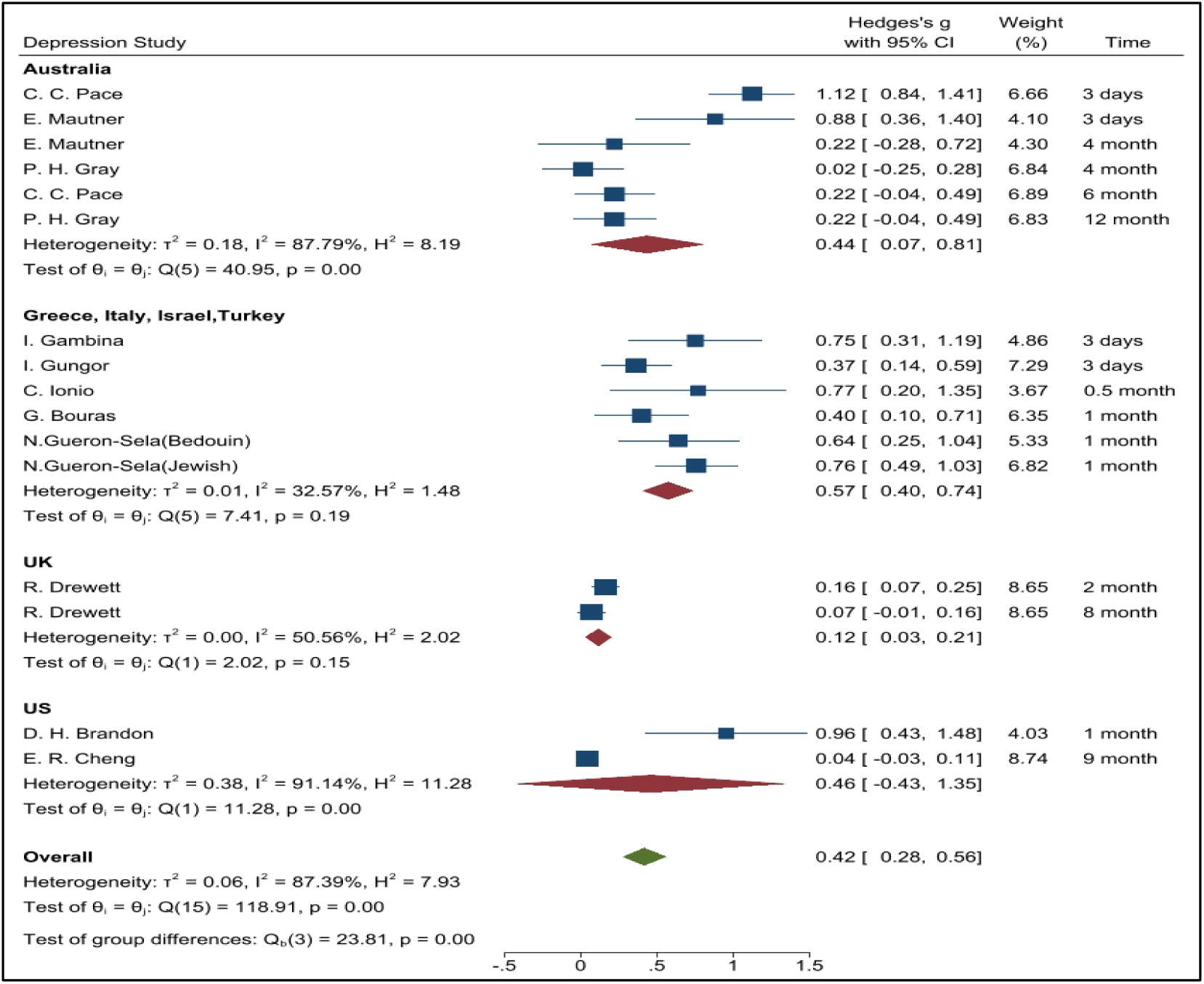
indicates the subgroup analysis conducted by region.

**Supplementary Figure 2.**
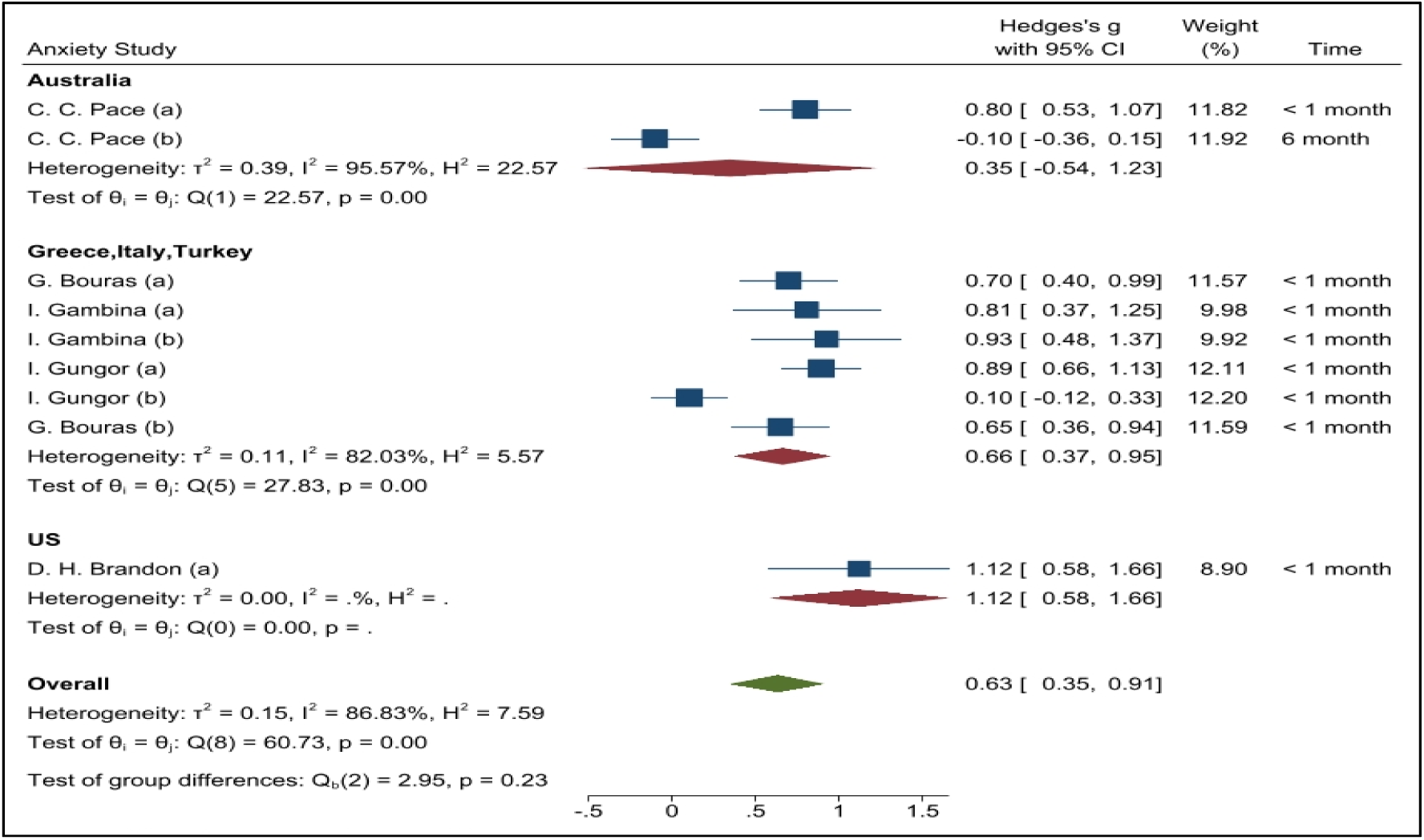
indicates the subgroup analysis conducted by region for anxiety.

**Supplementary Figure 3.**
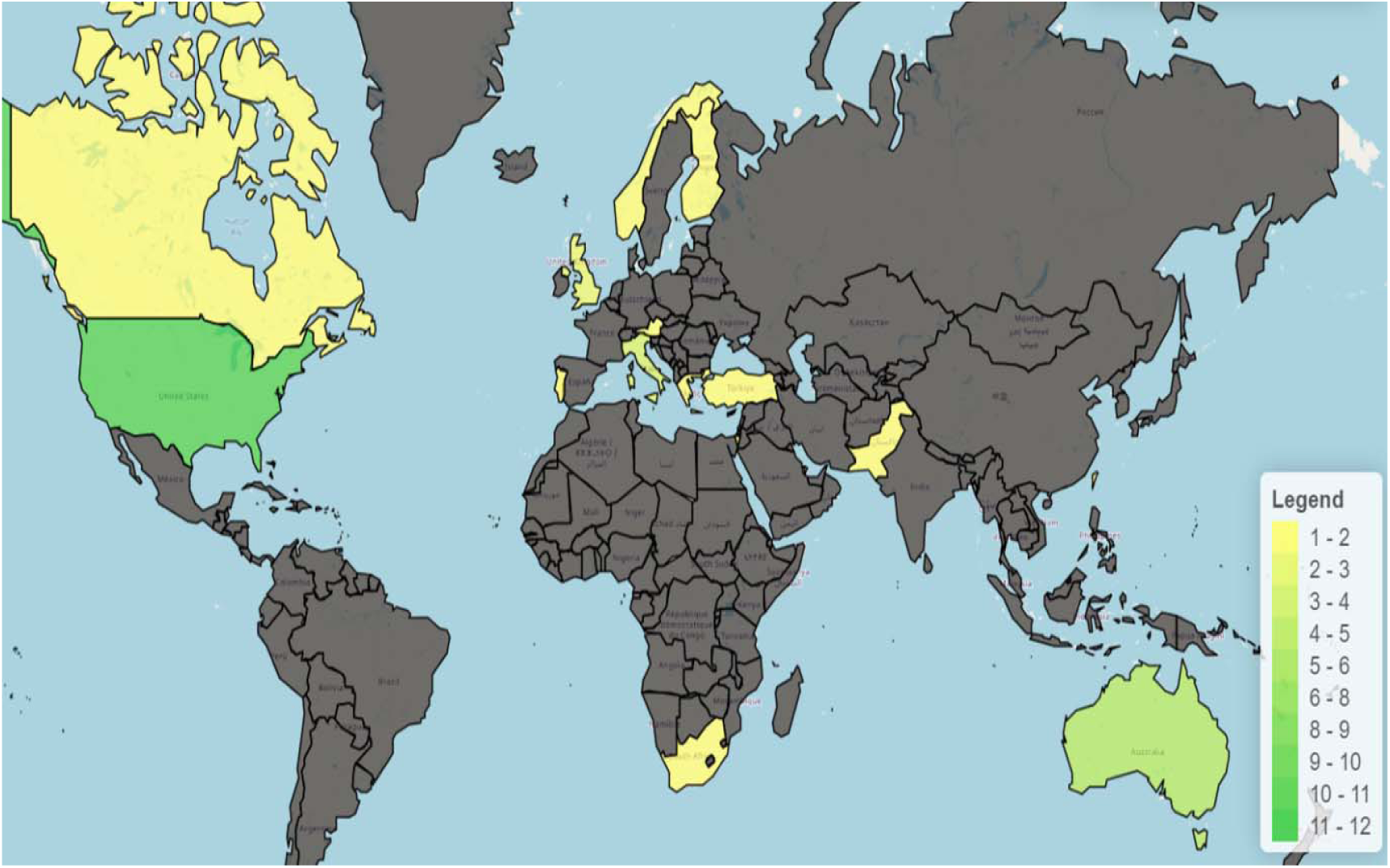
indicates the subgroup analysis conducted by outcome for stress.

**Supplementary Figure 4.**
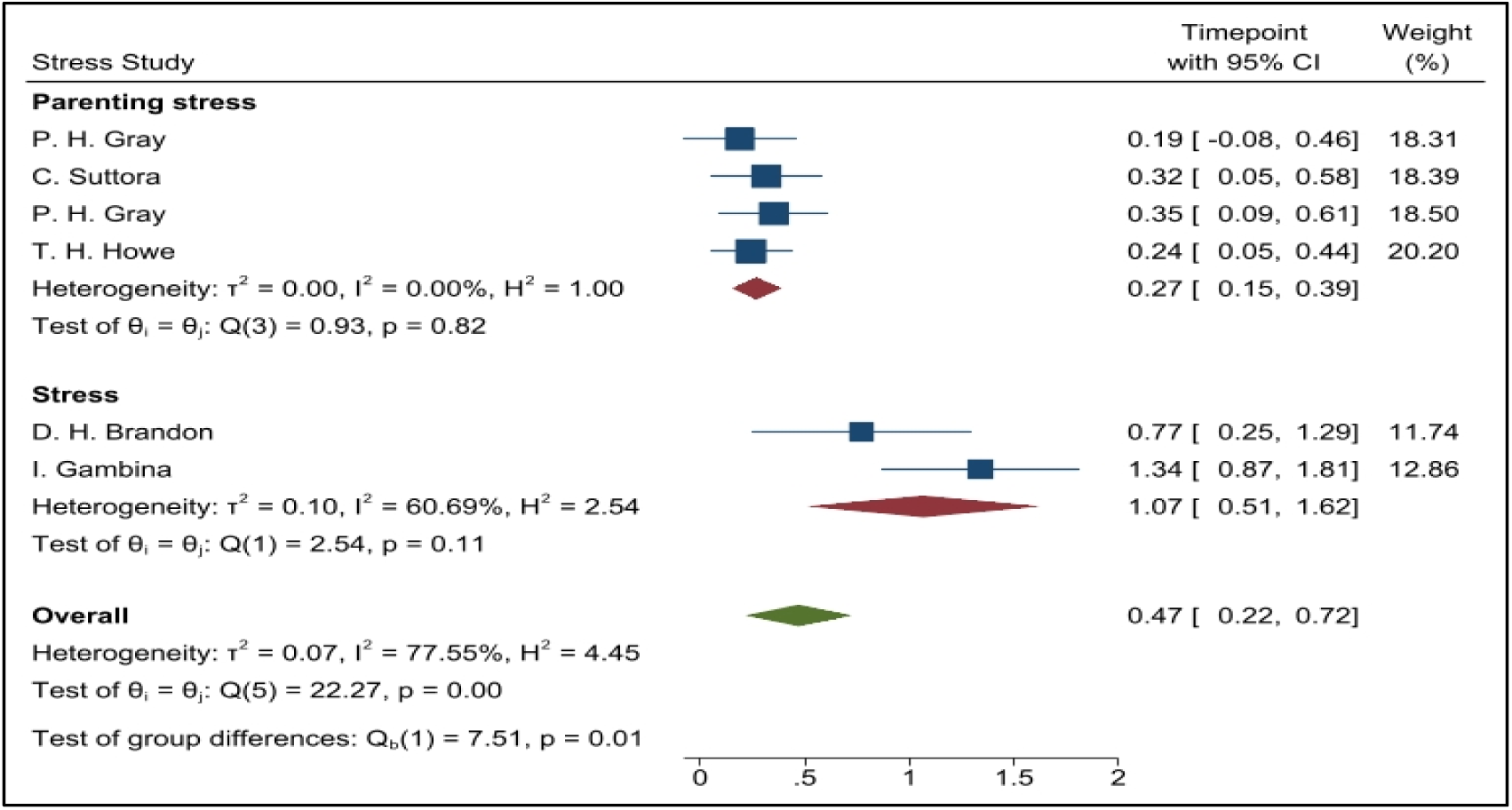
indicates the subgroup analysis conducted by outcome for stress.

**Supplementary Figure 5.**
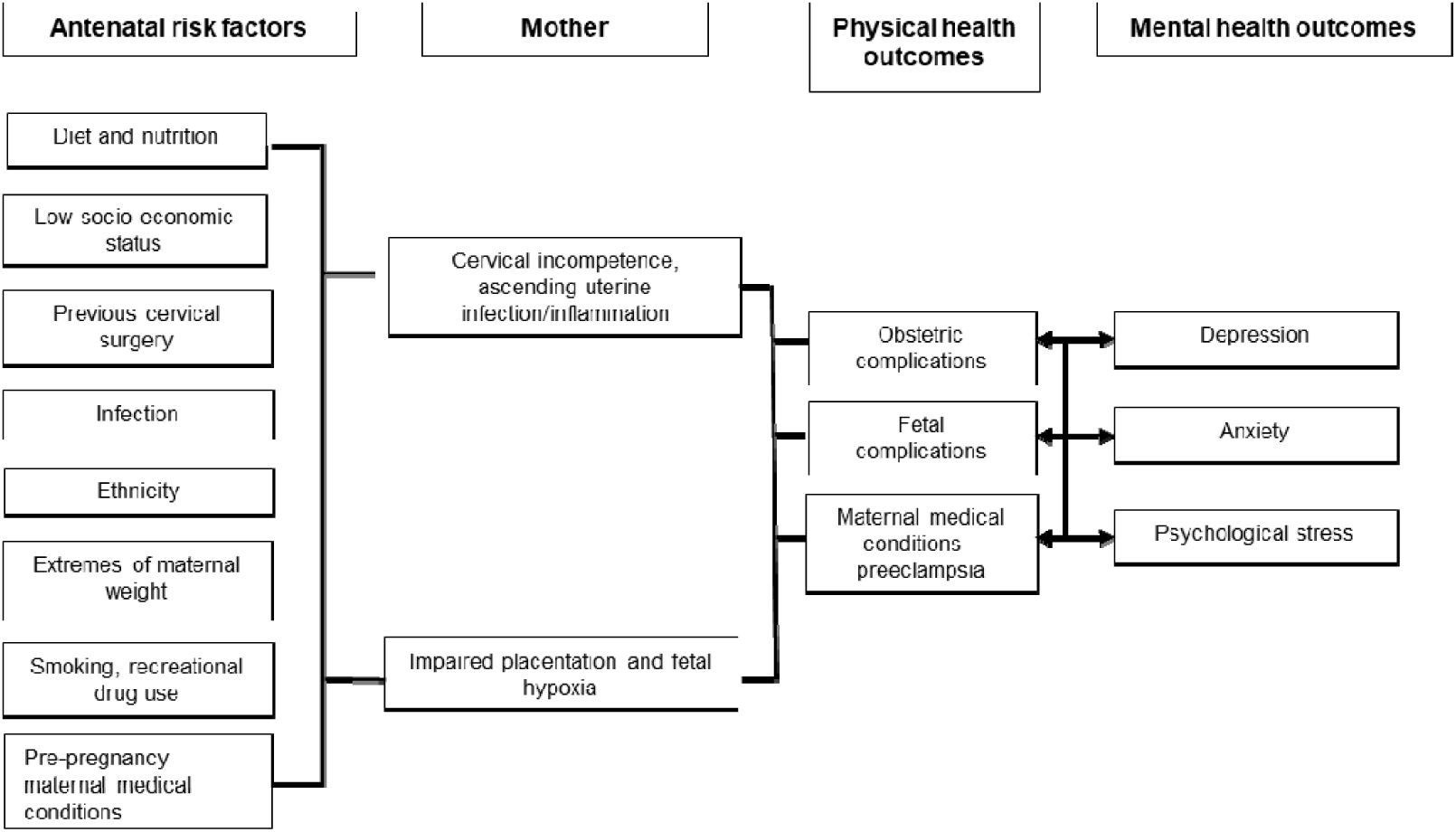
indicates a Preterm birth and Mental Health ‘Causation Tree’.

**Supplementary Figure 6.**
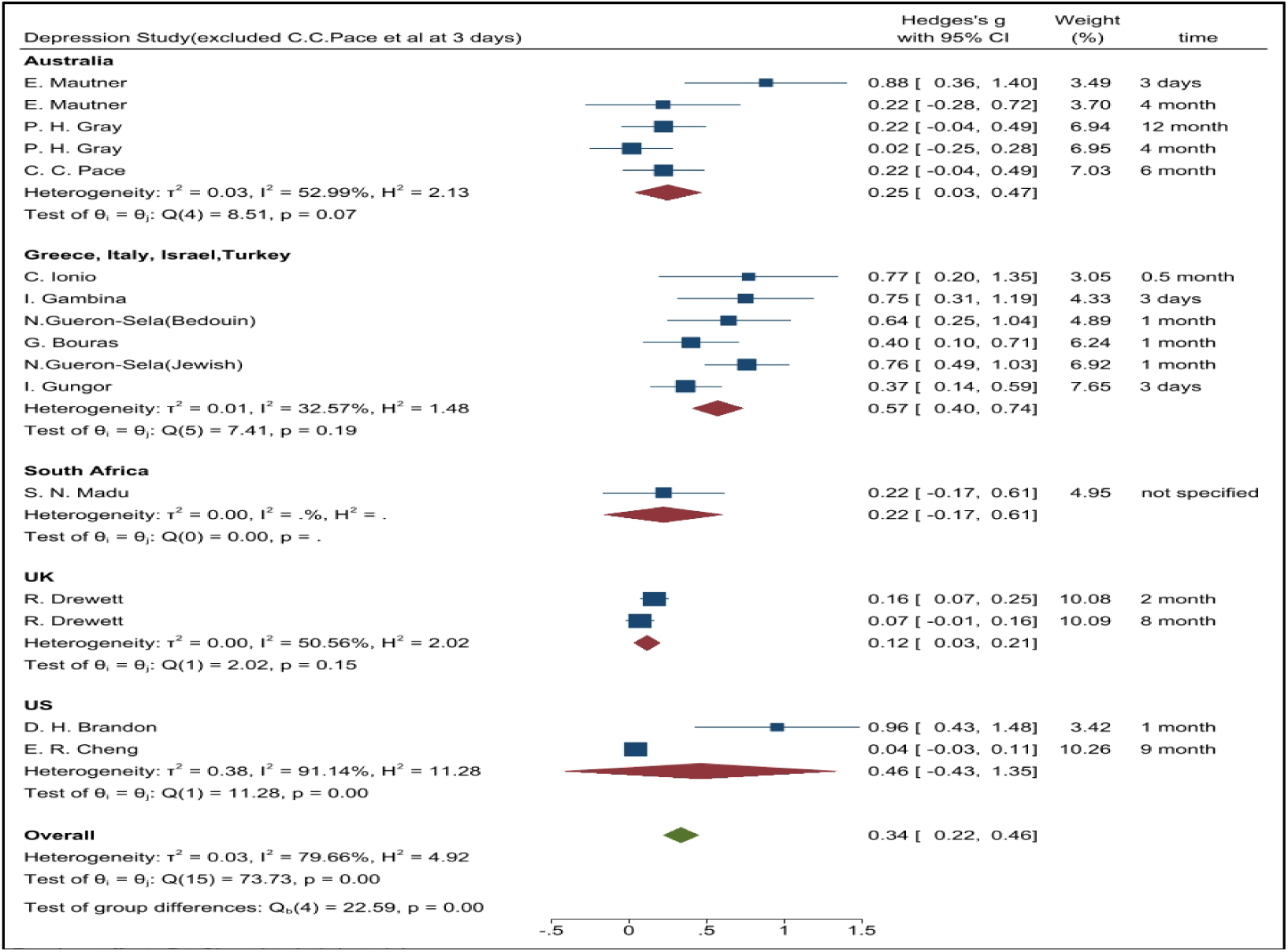
indicates the sensitivity analysis excluded one depression study (C.C.Pace et al [32])

**Supplementary Figure 7.**
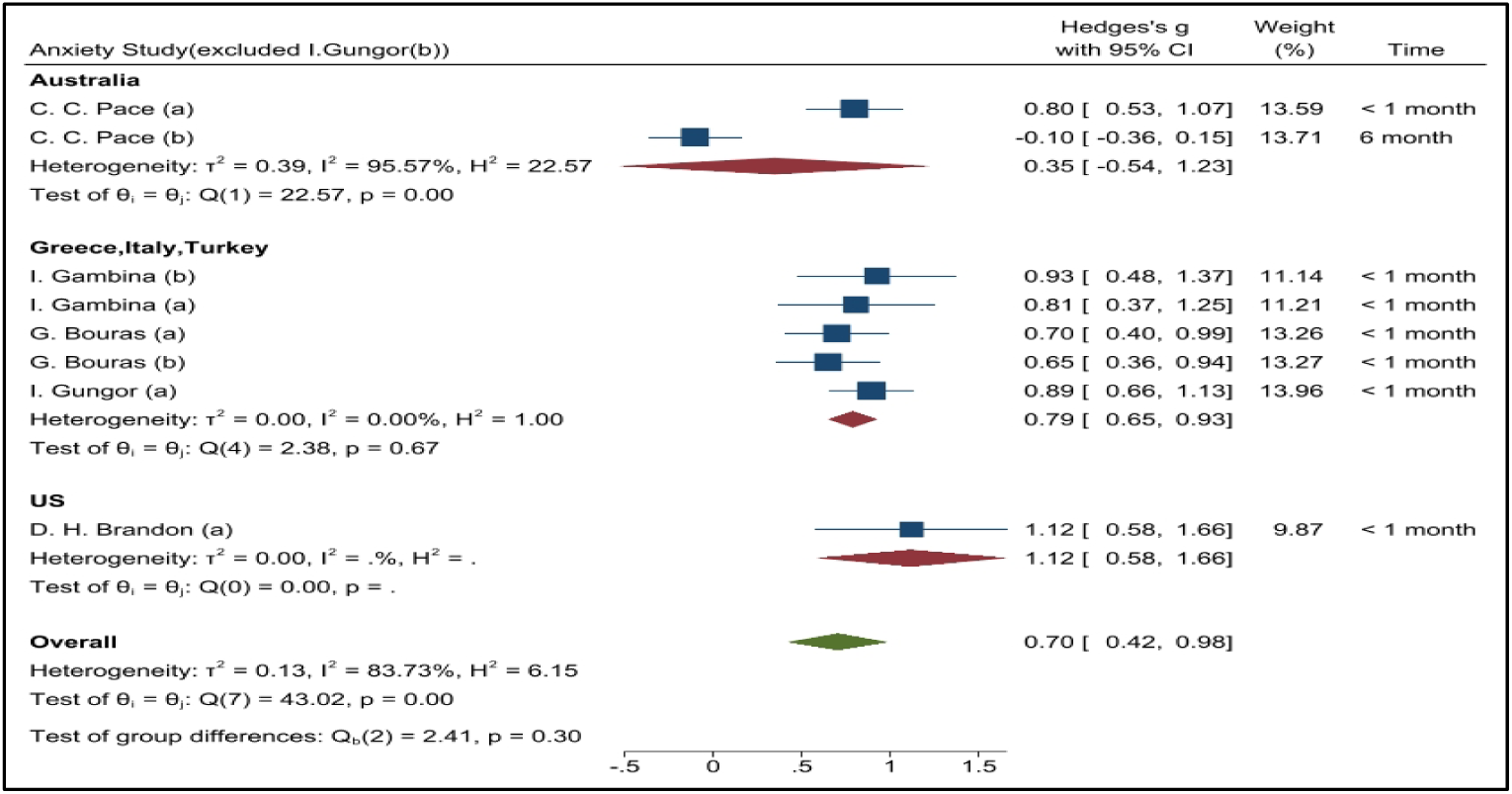
indicates the sensitivity analysis excluded one study (Gungor (b) et al [35])

**Supplementary Figure 8:**
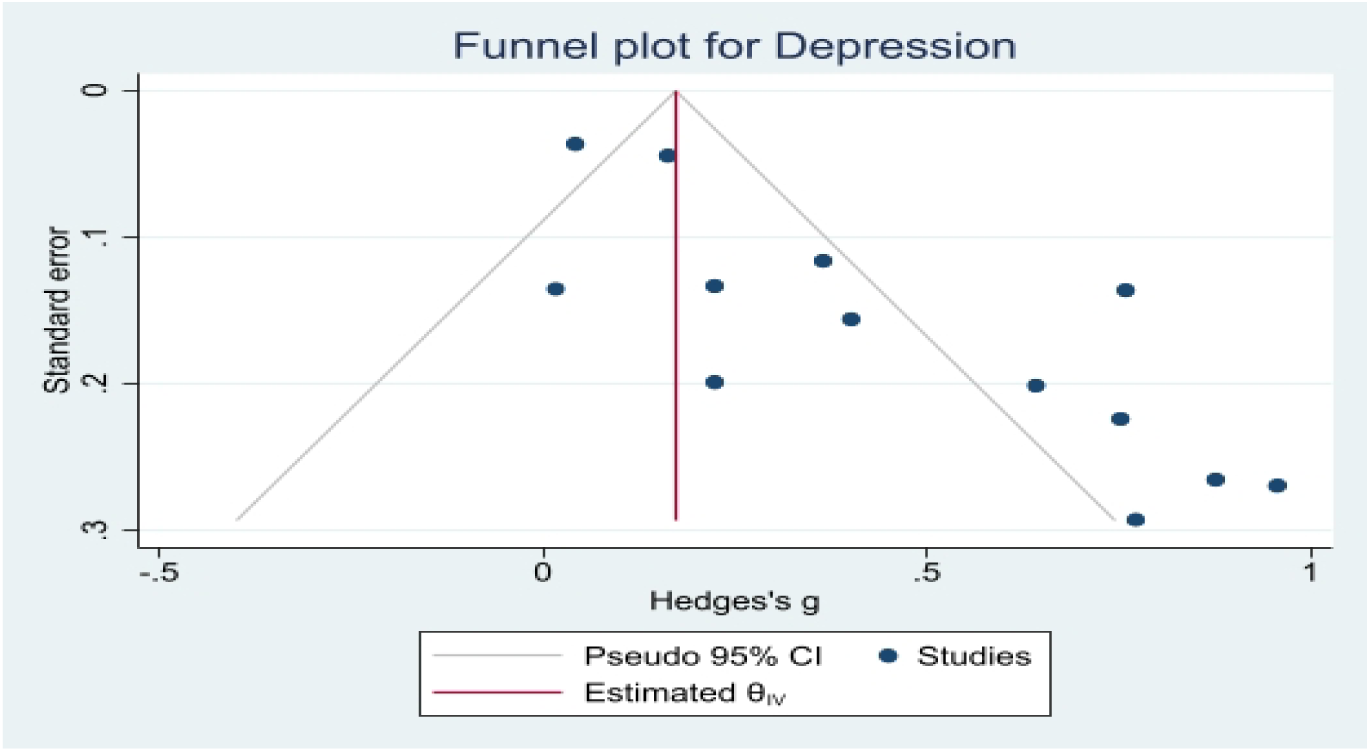
Funnel plot of studies for depression comparing full term vs preterm birth.

**Supplementary Figure 9:**
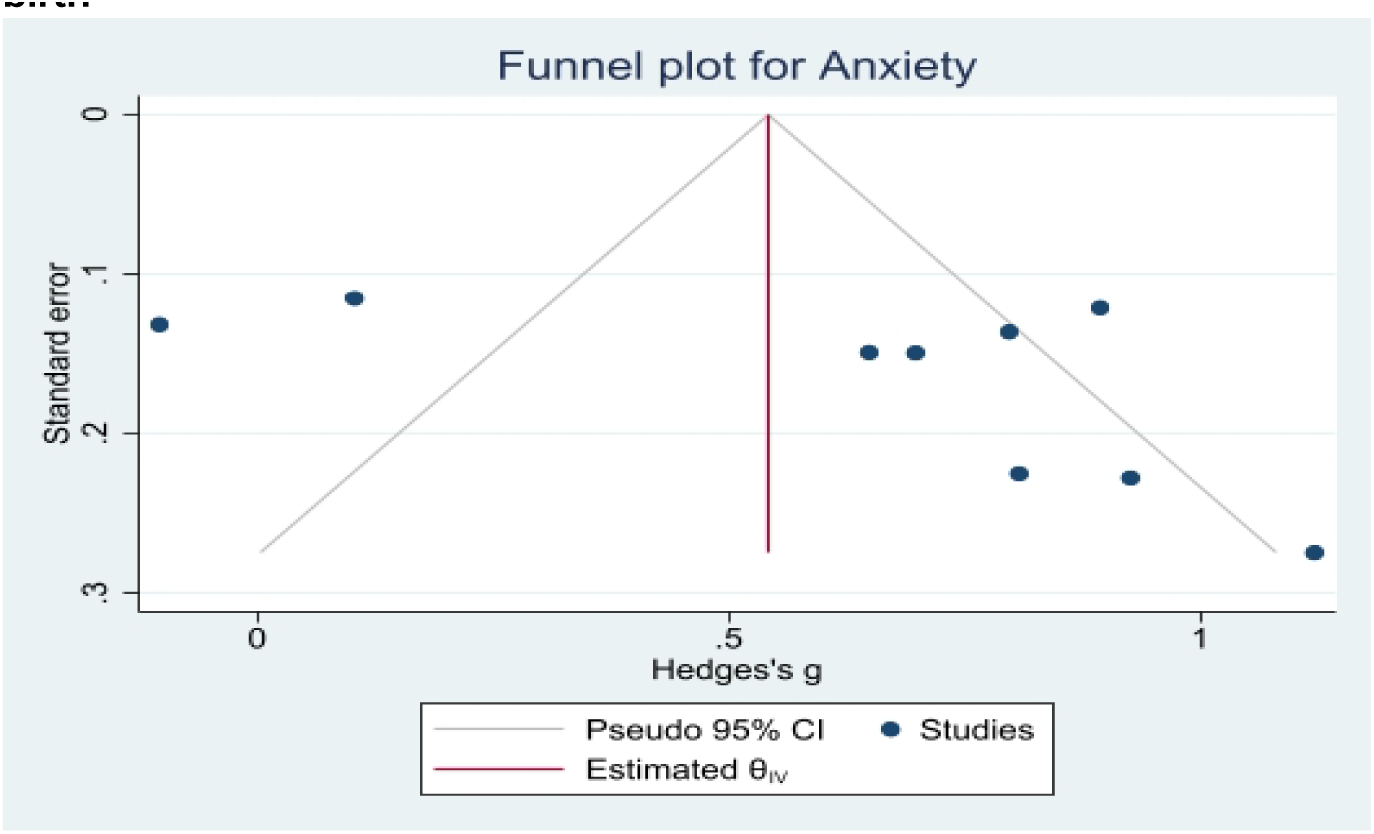
Funnel plot of studies for anxiety comparing full term vs preterm birth.

**Supplementary Figure 10:**
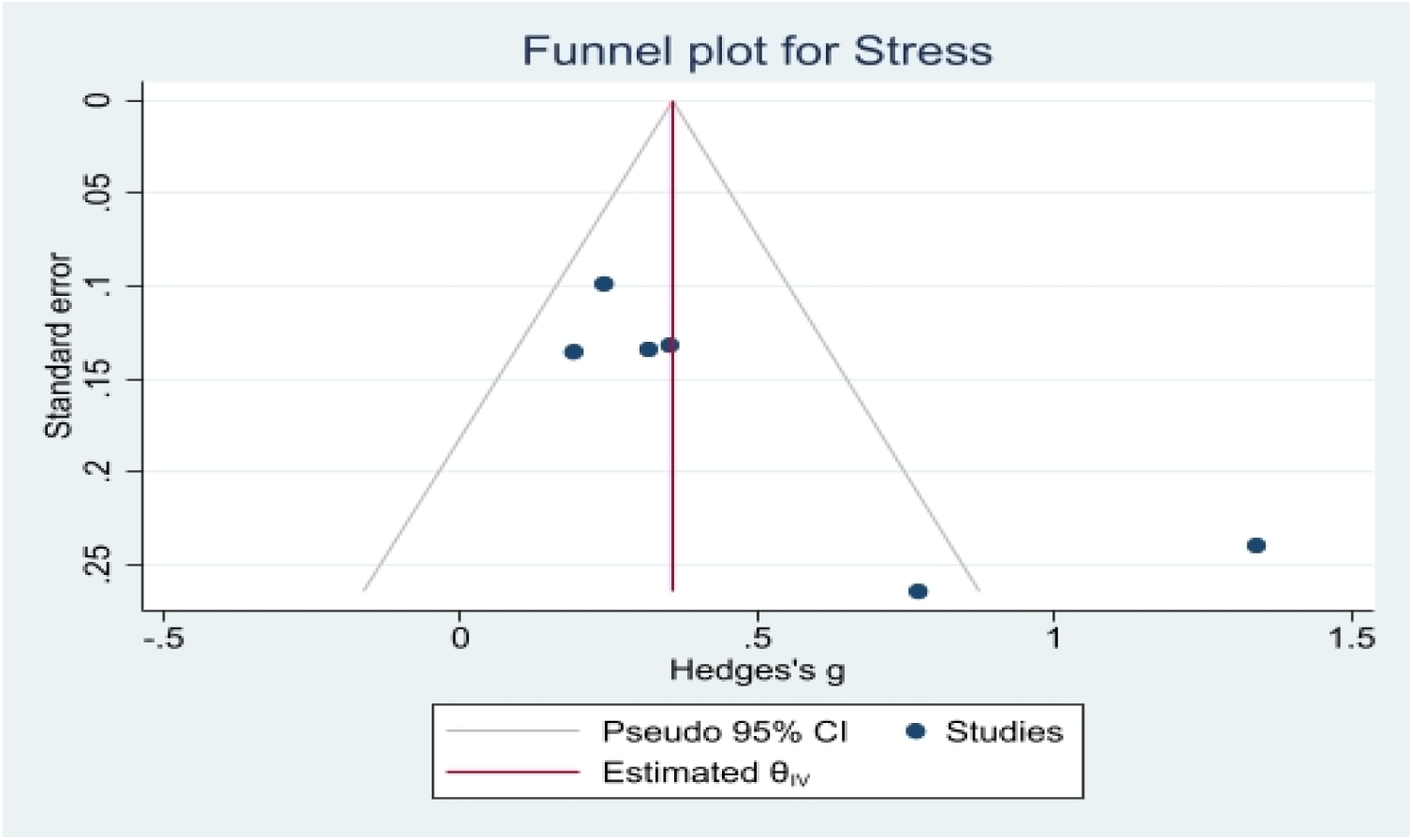
Funnel plot of studies for stress comparing full term vs preterm birth.

**Supplementary Figure 11.**
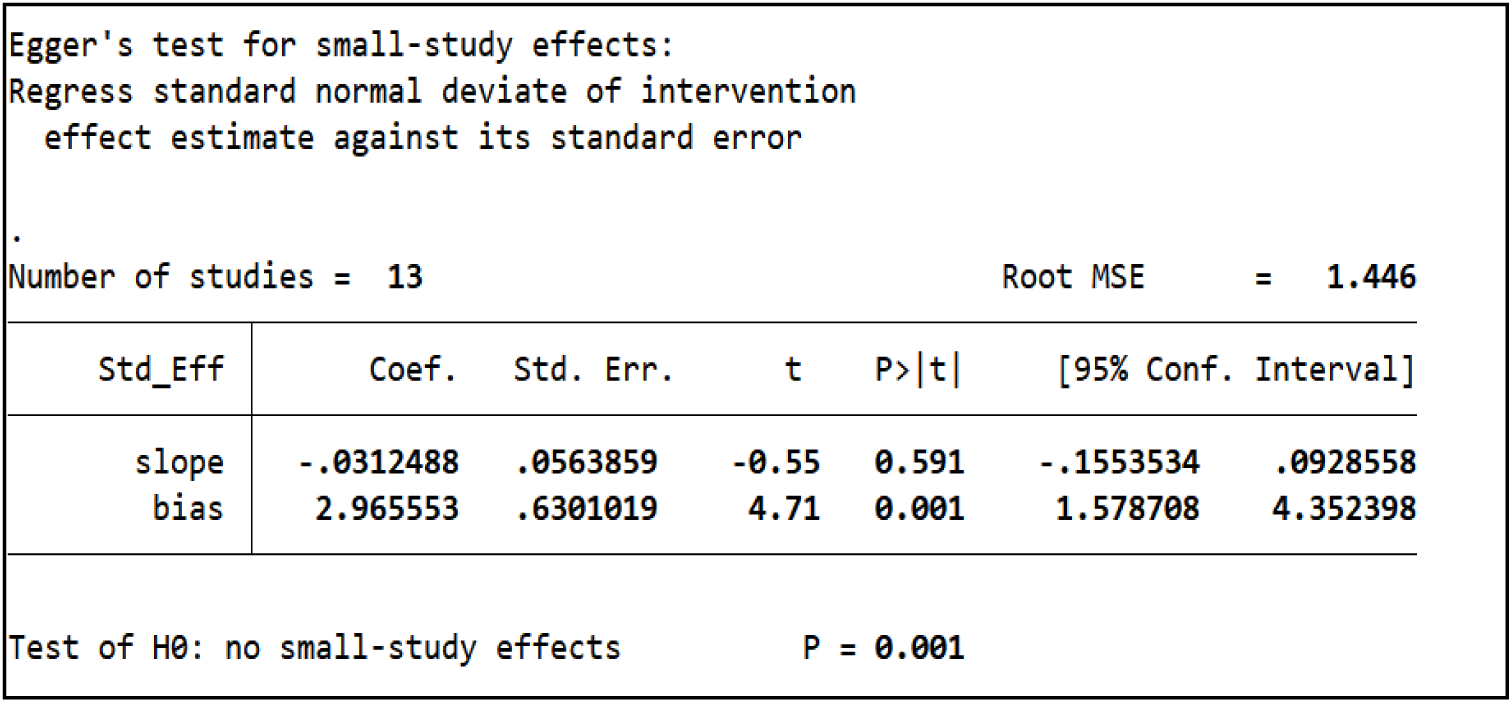
Egger’s test for the studies in meta-analysis for depression.

**Supplementary Figure 12.**
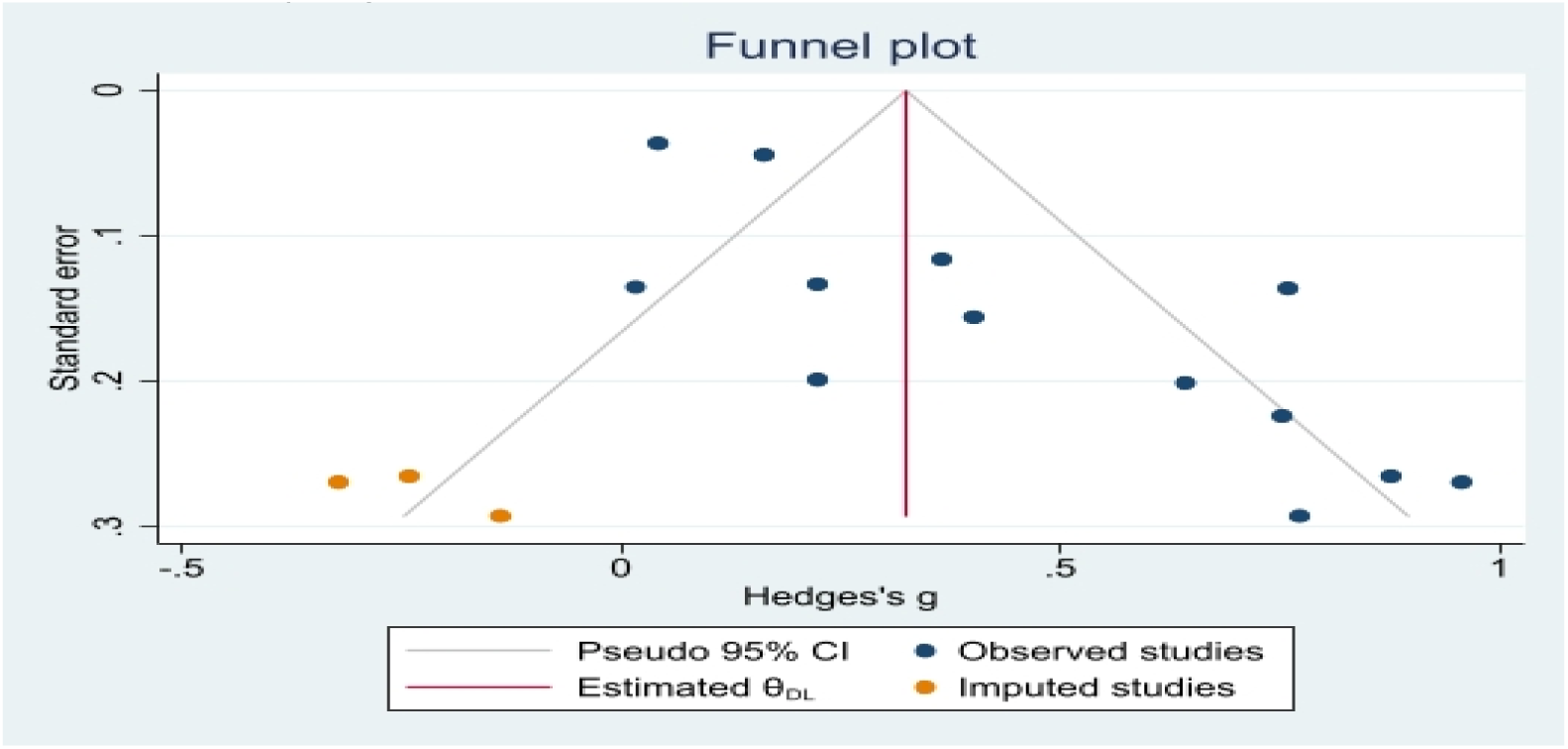
Trim and fill method to correct the effect of small studies.

**Supplementary Figure 13.**
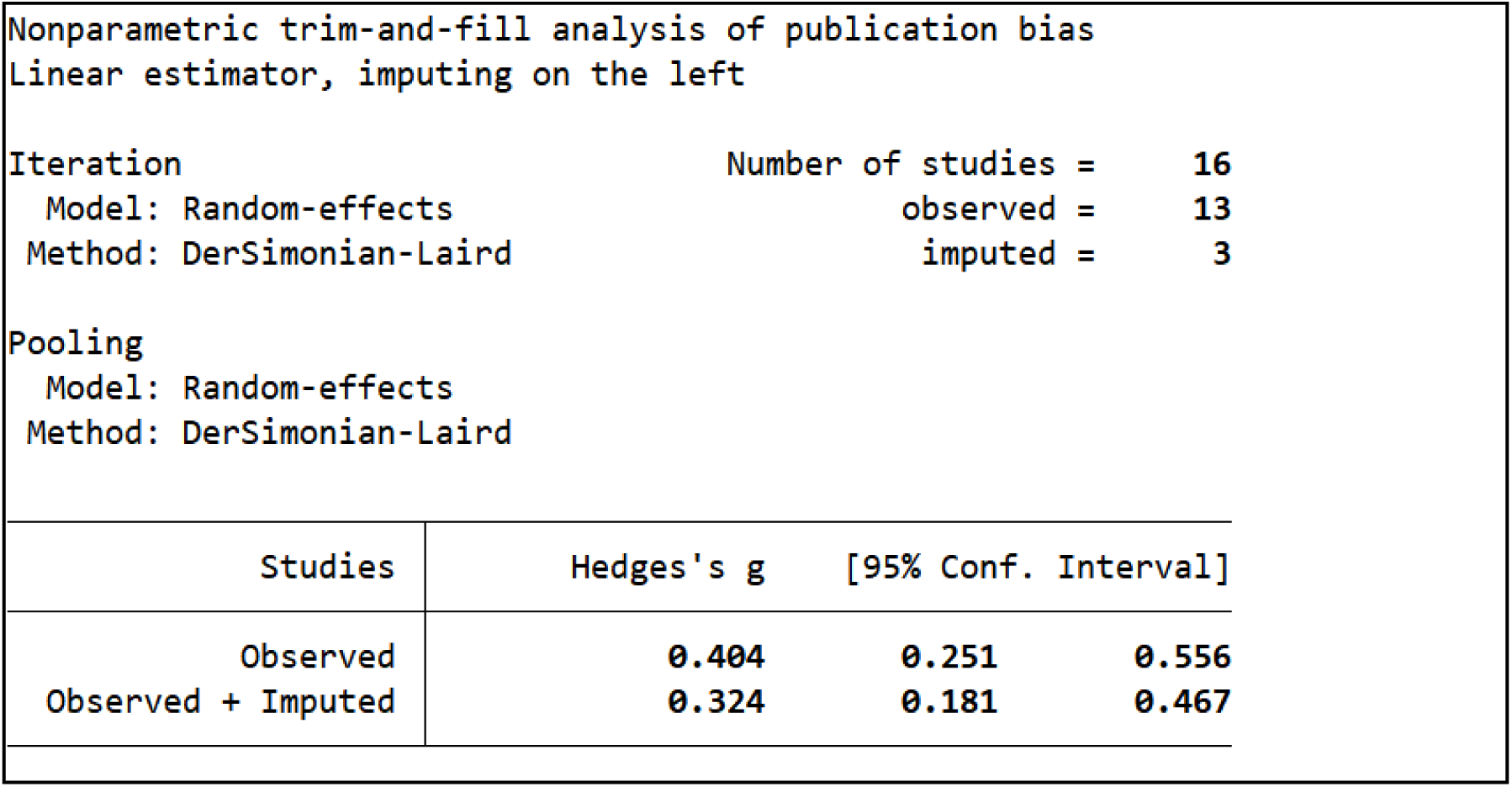
Statistic report of Trim and Fill Method.

**Supplementary Figure 14.**
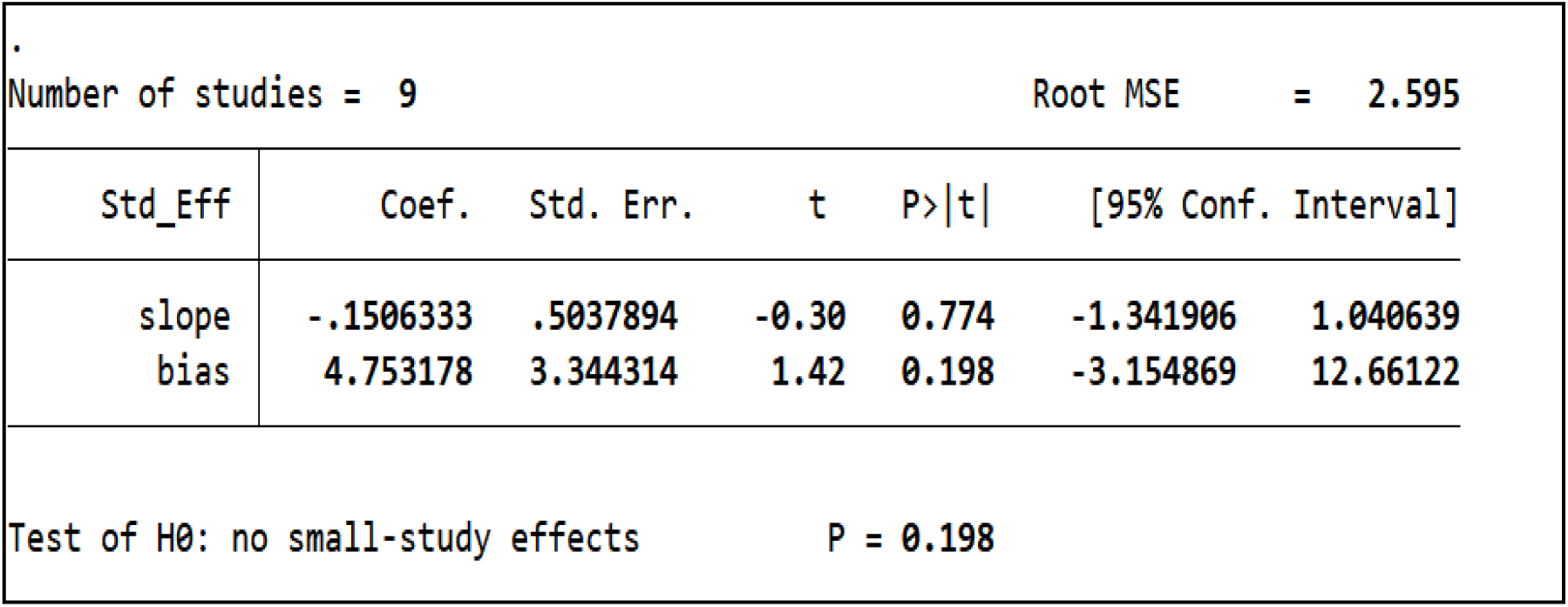
Egger’s test for the studies in meta-analysis for anxiety.

**Supplementary Figure 15.**
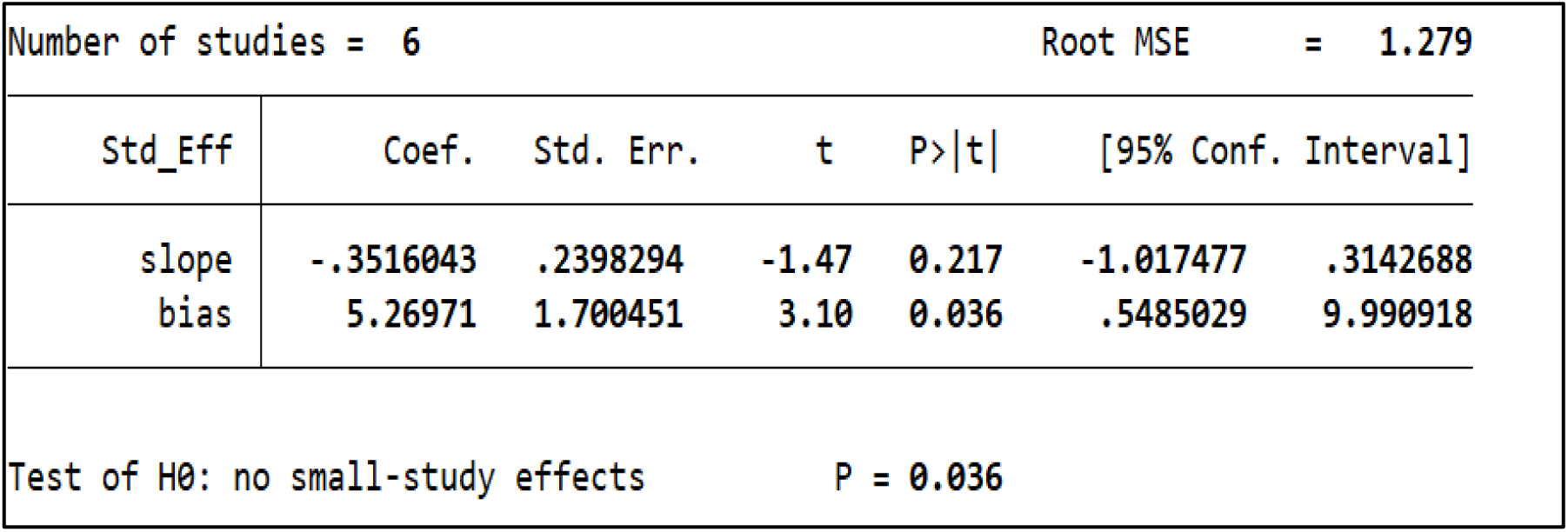
Egger’s test for the studies in meta-analysis for Stress.

